# Causal Analysis of Gynecologic Disorders and Adult-Onset Psychiatric Conditions: Integrating Matched Cohort, Polygenic Risk Score, and Mendelian Randomization

**DOI:** 10.64898/2026.11.12.26367938

**Authors:** Khaoula Ben Messaoud, Zeynep Asgel, Frederick Licciardi, Magdelena Janecka

## Abstract

**Background:** Endometriosis, polycystic ovary syndrome (PCOS), and uterine fibroids are linked to psychiatric morbidity, but evidence on causality across the full psychiatric spectrum remains limited.

**Methods:** We assessed associations between endometriosis, PCOS, uterine fibroids and adult- onset psychiatric conditions (schizophrenia and other psychotic disorders, mood disorders, anxiety and stress-related disorders) in UK Biobank female participants using three complementary approaches: a matched-pair cohort design, a polygenic risk score (PRS)-based cohort analysis, and bidirectional two-sample Mendelian randomization (MR). Sensitivity analyses were performed to test robustness.

**Results:** Endometriosis was associated with mood disorders (OR= 1.51, 95% CI 1.40, 1.63), anxiety and stress-related disorders (OR=1.49, 95% CI 1.38, 1.62), and behavioral syndromes with physical components (OR= 1.28, 95% CI 1.01, 1.62). Uterine fibroids were associated with mood disorders (OR=1.17; 95% CI 1.11, 1.23), anxiety and stress-related disorders (OR=1.40, 95% CI 1.34, 1.48), and behavioral syndromes (OR=1.29, 95% CI 1.12, 1.48). PCOS was associated with mood disorders (OR=1.47, 95% CI 1.32, 1.64) and anxiety and stress-related disorders (OR=1.79, 95% CI 1.62,1.99), but not behavioral syndromes. No gynecologic disorder was associated with schizophrenia or other psychotic disorders. Genetic analyses supported shared liability and causal effects between fibroids and anxiety disorders.

**Conclusions:** Women with gynecologic disorders are at increased risk of mood and anxiety- and stress-related disorders, and those with endometriosis or fibroids also face additional risk of behavioral syndromes. At least some of these risks appear driven by genetic liability, supporting routine mental health screening in gynecologic care and integrated treatment of gynecologic and mental health.

**Key Messages:**

- Women with endometriosis, polycystic ovary syndrome, or uterine fibroids have 17–80% higher risks of mood and anxiety/stress-related disorders but not psychotic disorders.
- Genetic analyses support shared genetic liability between uterine fibroids and psychiatric disorders, with evidence for a causal effect of fibroids on anxiety disorders.
- These findings support integrating routine mental health screening into gynecologic care for affected women.

## INTRODUCTION

Gynecologic disorders (GDs), including endometriosis, polycystic ovary syndrome (PCOS), and uterine fibroids (leiomyomas), are common conditions associated with substantial morbidity across the life-course. Beyond pain, fertility-related concerns, and reduced quality of life, these disorders may also be linked to psychiatric conditions. However, evidence remains incomplete, with most prior studies focusing on selected disorder pairs—particularly endometriosis and depression or anxiety—and often using short follow-up or heterogeneous clinical definitions.^1–3^

Because GDs are frequently underdiagnosed and diagnosed late, observational research is vulnerable to misclassification and residual bias.^4–7^ Genetically informed approaches may help to investigate those links and identify shared liability and potential causal effects,^8^ but such studies remain limited or inconsistent and have rarely examined uterine fibroids or the full range of adult- onset psychiatric disorders.^3,9–14^ Supplemental Figure 1 summarizes the literature on GDs and adult-onset psychiatric conditions.

We therefore investigated associations between endometriosis, PCOS, uterine fibroids, and adult- onset psychiatric conditions in the UK Biobank (UKB) using 3 complementary approaches: matched-pair analyses, polygenic risk score (PRS) analyses, and bidirectional two-sample Mendelian randomization (MR). We assessed observational associations, individual-level genetic overlap, and the direction of potential causal effects, with sensitivity analyses to test robustness.

## METHODS

### Data source and study population

The UKB is a prospective cohort of more than 500,000 UK participants aged 40–69 years at recruitment (2006–2010), with linked hospital, primary care, self-report, and genetic data. We included all female participants and identified first recorded diagnoses of gynecologic and psychiatric disorders through December 31, 2024. The cohort profile is detailed elsewhere.^15^ Hospitalization data were available for all participants (100%), while primary care data were available for 45% of the UKB cohort. All females in the UKB cohort were included in our study.

### Exposure: Gynecologic disorders

#### Diagnoses of gynecologic disorders

We investigated GDs, including endometriosis, PCOS, and uterine fibroids. These GD diagnoses were identified using first recorded diagnosis codes in Table 1.

**Table 1:**
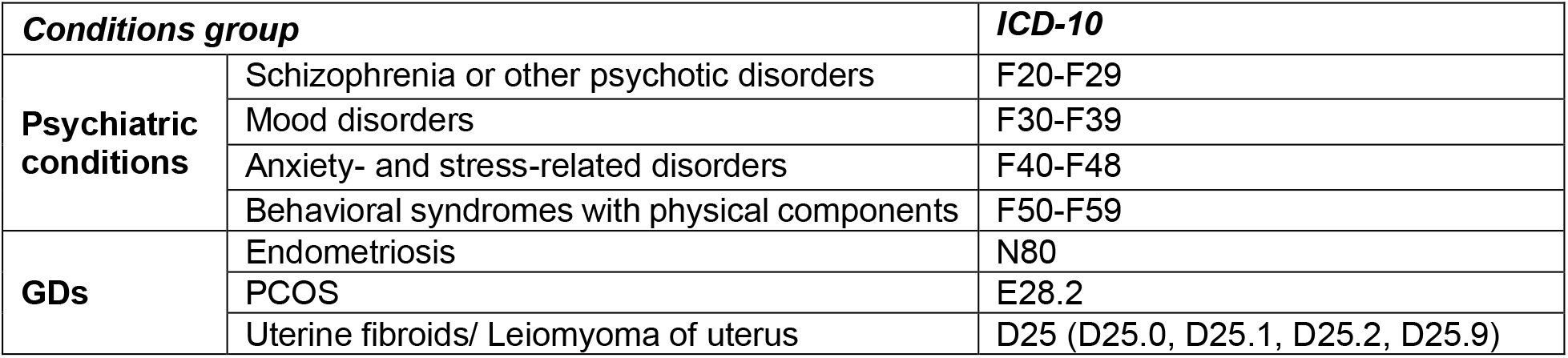
Psychiatric conditions, GDs and related ICD-10 code ranges.

#### Polygenic risk scores and genetic instruments for GDs

We used large-scale GWAS summary statistics for endometriosis, PCOS, and uterine fibroids,^16–18^ harmonized to the GRCh37/hg19 build. We calculated PRS to quantify individual-level genetic liability to each GD using SNP weights from these GWAS. The same GWAS summary statistics were also used to select genetic variants as instrumental variables for two-sample MR (Appendix Material S1; summary statistics in Supplemental Table S1).

### Outcome: Psychiatric disorders

Outcomes were first diagnoses of schizophrenia and other psychotic disorders, mood disorders, anxiety- and stress-related disorders, and behavioral syndromes with physical components, defined using ICD-9/10 codes (Table 1).

### Covariates

Socio-demographic variables were ascertained at the time of enrollment: year of birth, self- reported race, education, household income, area-level Index of Multiple Deprivation (IMD), and migration status.

**Race** was categorized as White (ref), more than one race, Asian or Asian British, and Black or Black British.

**Education** was grouped as: college or university degree; Certificate of Secondary Education (CSE); other professional qualifications; Advanced Level (A/AS) levels (ref); Ordinary Level or General Certificate of Secondary Education (O/GCSE) levels; National Vocational Qualification (NVQ), Higher National Diploma (HND), or Higher National Certificate (HNC); and “none of the above.”

**Household Income** (pre-tax) was categorized as <£18,000 (ref), £18,000–30,999, £31,000– 51,999, £52,000–100,000, >£100,000, and “prefer not to answer”.

The **IMD** was assigned based on residential address and analyzed in quintiles within each local council in England, Scotland, and Wales (first quintile = least deprived, ref)

**Migration status** was defined as being born outside the UK (yes/no (ref)).

We accounted for the presence of potential metabolic/endocrine confounders (yes/no (ref)). These conditions included obesity (E65–E68), diabetes (E10–E13), thyroid disorders (E00–E07), or other metabolic disorders (E70–E80, E83–E88). We only controlled for metabolic/endocrine conditions diagnosed prior to the first occurrence of the studied RDs diagnosis, as conditions occurring after the RD diagnosis may lie on the causal/functional pathway between reproductive and psychiatric conditions.

**Reproductive disorder** co-occurrence (yes/no (ref)) was defined as another studied RD diagnosed before the index reproductive diagnosis.

### Study design

We combined three analytic designs to provide complementary insights into the relationships between GDs and psychiatric conditions. First, to examine observational associations between GDs and psychiatric outcomes, we used matched-pair designs within the UKB, matching on primary care linkage status, year of birth, and IMD quintile; non-diagnosed participants had no record of the specific GD in hospital or primary care data, and matching ratios were 1:1 for endometriosis and fibroids and 1:5 for PCOS, chosen to enable inclusion of all cases in the UKB (higher matching ratios for endometriosis and PCOS would require case exclusion). Second, we implemented PRS-based analyses of genetic liability for GDs and psychiatric conditions. Third, to evaluate the directionality of potential causal associations, we applied bidirectional two-sample MR.

### Statistical analysis

The hypothesized causal framework is shown in Supplemental Figure S2.

### Observational analysis

We used multivariable conditional logistic regression in the matched-pair designs to estimate odds ratios (ORs) and 95% confidence intervals (CIs) for associations between gynecologic diagnoses and psychiatric outcomes. We examined two periods: We examined two periods: (i) the full study period (from 18 years before to 30 years after the index date) and (ii) after the GD diagnosis (0– 30 years). To ensure robust effect estimation, we required at least 5 psychiatric outcome cases among women with GDs. All models were adjusted for primary care data availability, IMD, and year of birth, with stepwise inclusion of socio-demographic variables (race, education, income, migration status), and metabolic/endocrine conditions.

### Sensitivity in observational analysis

To account for co-occurring GDs, we (1) included all GDs as additional predictors in the same model; (2) examined the associations between GDs and psychiatric conditions after excluding women with multiple GDs and controls with any of the investigated GDs from the sampling pool. To optimize the matching ratios in this sensitivity analysis, we used ratios of 1:6 for endometriosis, 1:12 for PCOS, and 1:2 for fibroids. Furthermore, to ensure the consistency of our findings across primary care linkage status, we (3) replicated the observational analysis in the participants with primary care data linkage using a matching ratio of 1:1 for endometriosis and PCOS, and 1:5 for fibroids.

### PRS analysis

We investigated associations between PRSs for each GD (endometriosis, PCOS and uterine fibroids),^19–21^ and the risk of lifetime psychiatric diagnosis. For each psychiatric outcome, we performed a logistic regression with the standardized PRS as exposure. Models were adjusted for the first 10 genetic principal components to account for population structure, and we reported odds ratios (ORs) with 95% confidence intervals (CIs). Details of PRS construction are provided in Supplemental Section S1.

### Mendelian Randomization

We performed bidirectional two-sample MR analyses using the TwoSampleMR^22^ and ieugwasr R packages, treating each GD and psychiatric outcome in turn as both exposure and outcome to assess directionality and potential reverse causality (using the GWAS described above). The primary estimator was the inverse-variance weighted (IVW) method. Robustness was assessed using MR-Egger regression, weighted median, and weighted mode estimators. An MR heterogeneity test was performed to assess whether the genetic variants had consistent effects on the outcomes. To evaluate potential pleiotropy, we performed the MR-Egger intercept test for directional horizontal pleiotropy to determine whether any genetic variants directly influenced the outcomes independently of their indirect associations with the corresponding exposures in each MR direction. Furthermore, we carried out a leave-one-out analysis to test the stability of the findings by systematically removing each genetic variant one at a time, examining the impact on the overall results. Full instrument selection and harmonization details are provided in Supplemental section S2.

## RESULTS

### Descriptive analysis

We analyzed three matched case-control cohorts for endometriosis, polycystic ovary syndrome (PCOS), and uterine fibroids, with follow-up through 31 December 2024. The endometriosis cohort included 10 755 diagnosed women matched 1:1 to women without a diagnosis (n=21 510). The PCOS cohort comprised 2536 cases matched 1:5 to women without a diagnosis (n=17 752). The fibroids cohort included 26 057 cases matched 1:1 to controls (n=52 114). Cohort characteristics, including psychiatric diagnoses, sociodemographic factors, metabolic conditions, age at first diagnosis of the GD, and distributions of co-occurring GDs, are shown in Supplemental tables S2-S4.

### Observational analysis

Fully adjusted models showed that women with any of the GDs we studied had higher lifetime risks of mood and anxiety- and stress-related disorders (endometriosis odds ratio 1.51; 95% confidence interval 1.40-1.63; PCOS 1.47; 1.32-1.64; uterine fibroids 1.17; 1.11-1.23). The risks of anxiety and stress-related disorders were highest in women with PCOS (1.79; 1.62-1.99), followed by endometriosis (1.49; 1.38-1.62), and uterine fibroids (1.40; 1.34-1.48). The risks of behavioral syndromes with physical components were also higher in women with uterine fibroids (1.29; 1.12-1.48) and endometriosis (1.28; 1.01-1.62) but not with PCOS (1.31; 1.00-1.70) (Supplemental tables S5-S7 and figure 1).

**Figure 1:**
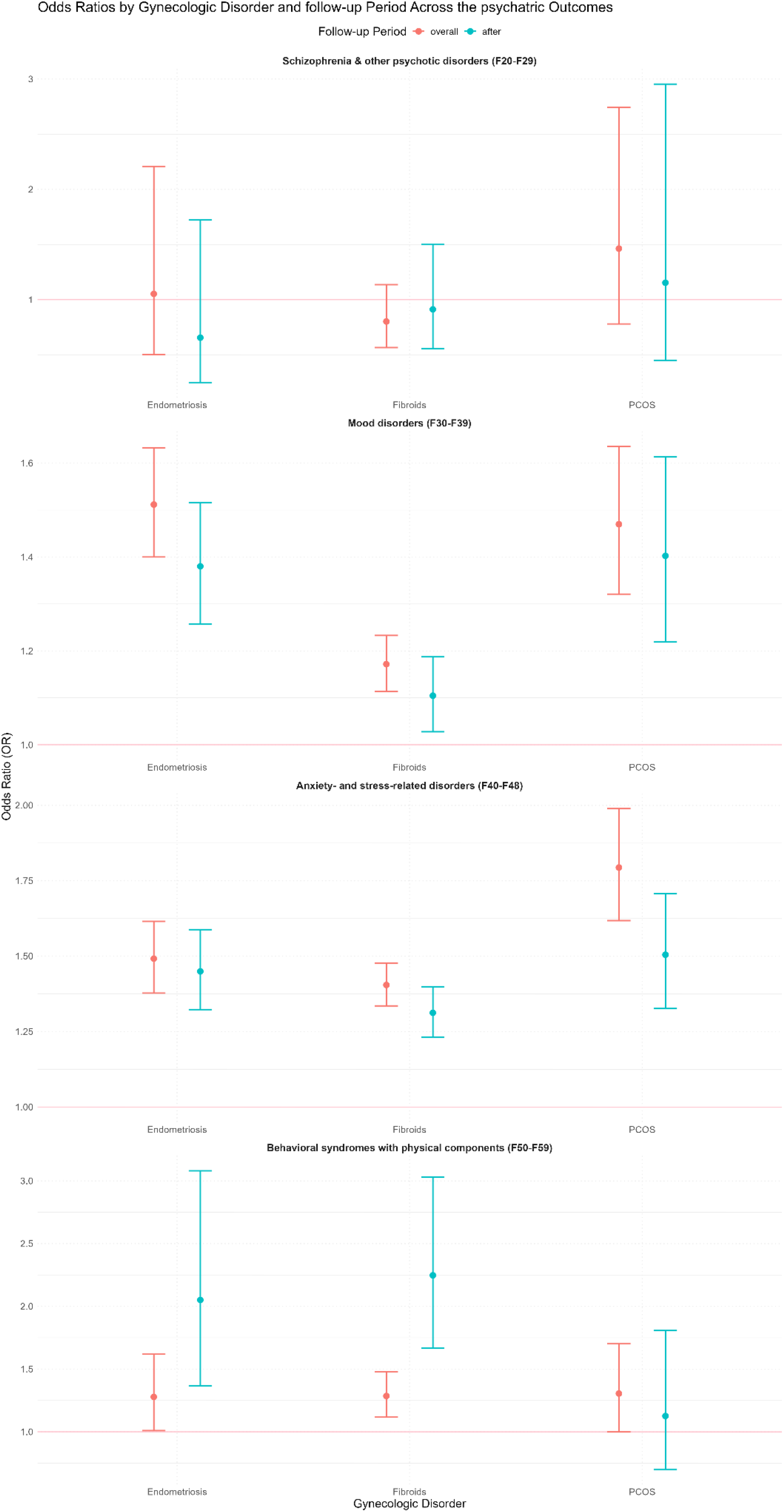
Association Between Endometriosis, PCOS, Fibroids and Psychiatric Outcomes by Time Period.

When restricted to the period after GD diagnosis, results were consistent with the lifetime analyses, with slightly lower risk estimates and larger confidence intervals (figure 1). However, the association between endometriosis, fibroids, and behavioral syndromes with physical components for the restricted period after the diagnosis showed a higher estimated risks and larger confidence intervals. The association between uterine fibroids and behavioral syndromes with physical components across the two follow-up periods was notably different, with no overlap between the confidence intervals in those analyses (1.29; 1.12-1.48 in the overall period vs 2.25; 1.67-3.03 in the after period) (Supplemental table S7). The association between PCOS and behavioral syndromes with physical components was still not significant for the period after the diagnosis (1.13; 0.70-1.81). No GD was associated with schizophrenia or other psychotic disorders in either period (Figure 1). No significant changes were observed in the association estimates between GDs and psychiatric conditions across the incremental adjustments for matching, sociodemographic variables, and metabolic/endocrine status in both periods of follow- up (Supplemental Tables S5, S6, and S7).

### Sensitivity analysis

Estimates adjusted for co-occurring GDs, estimates using strict case/control definitions, and analyses limited to participants with primary care linkage were consistent with the estimates from the main analyses. For all significant associations, confidence intervals were narrower in the primary care subgroup, indicating greater precision (Figure 2 and Supplemental Table S8).

**Figure 2:**
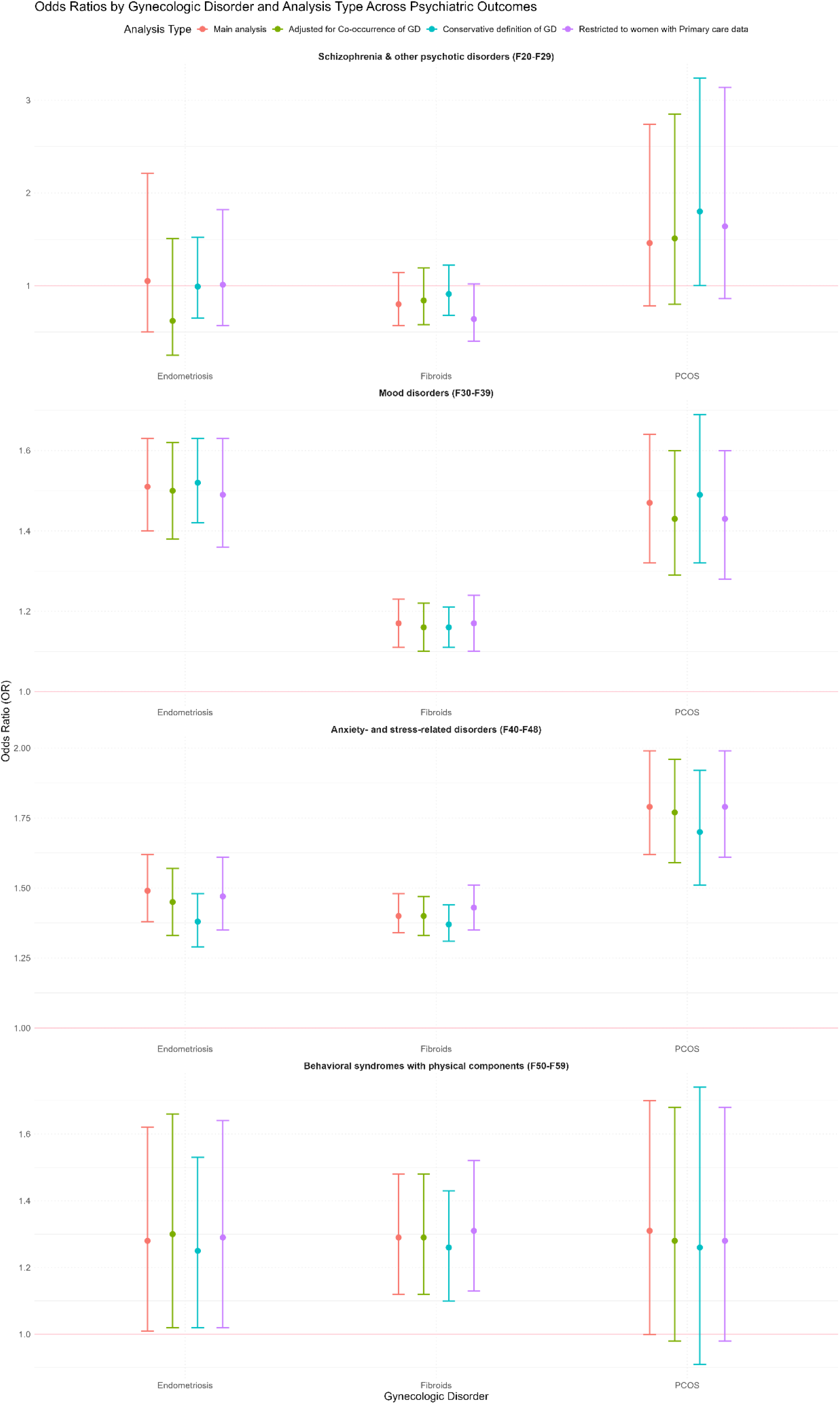
Sensitivity analysis of the Association Between Gynecologic disorders and Psychiatric Outcomes in the overall study Period.

### Polygenic Risk Score analyses

In PRS analyses, higher genetic liability to uterine fibroids was associated with mood and anxiety/stress-related disorders. Higher PRSs for endometriosis and PCOS were associated with small elevations in the odds of mood disorders, but these associations were not significant. No clear PRS associations were observed for psychotic disorders or behavioral syndromes. The detailed results are shown in Figure 3 and Supplemental Table S9.

**Figure 3:**
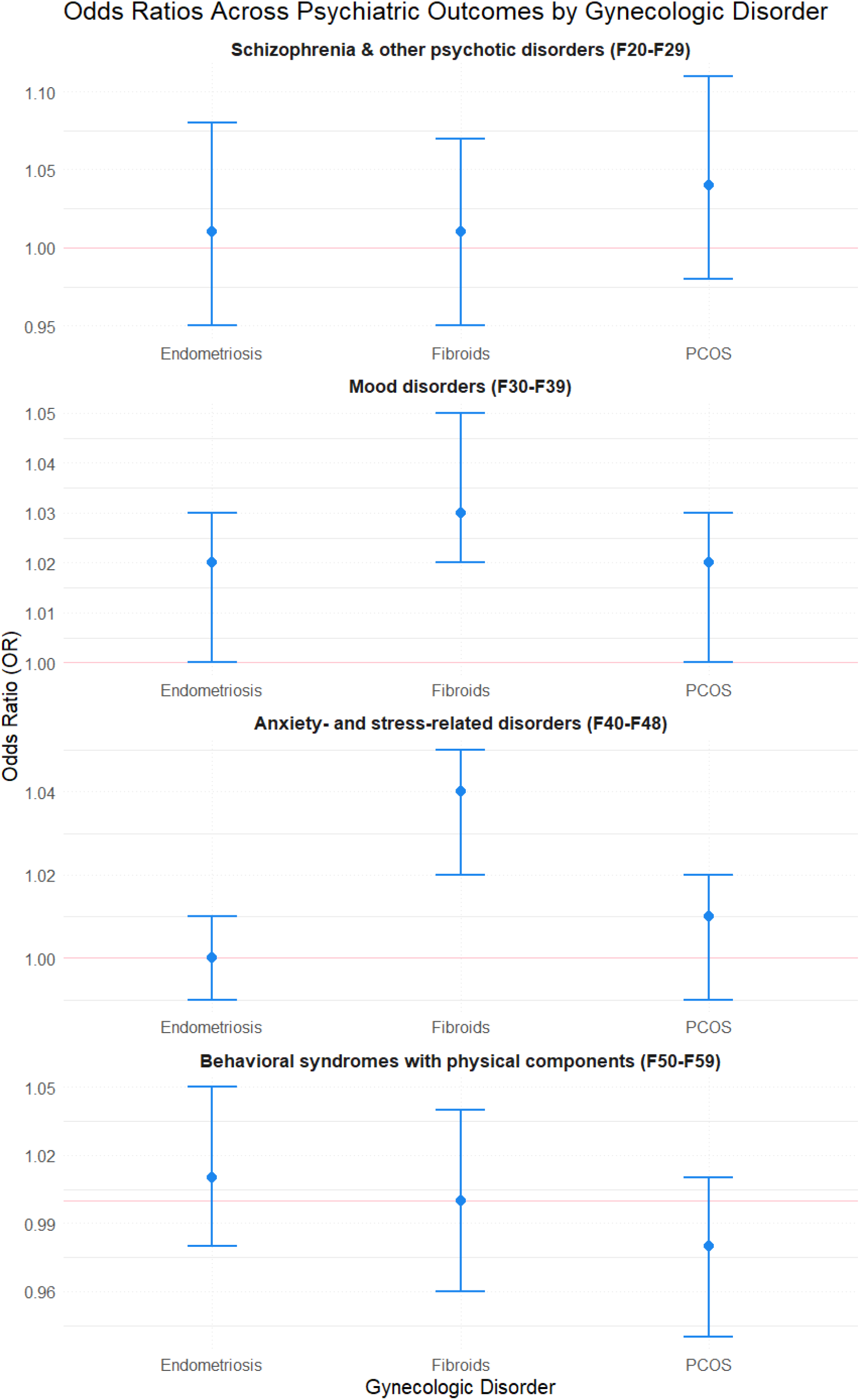
Analysis of the Association Between Gynecologic disorder and Psychiatric Outcomes in the overall study Period using PRS based definition of Gynecologic disorders.

### Mendelian Randomization

In bidirectional two-sample MR, genetic liability to uterine fibroids was associated with anxiety/stress-related disorders in the primary IVW analysis (Supplemental Table S10). This association was supported by MR-Egger and weighted median estimates, with a consistent direction of effect and similar magnitude (Supplemental Table S11). No other GD–psychiatric outcome pairs showed clear evidence of a causal association in the MR analyses. For the fibroid– anxiety/stress association, heterogeneity tests did not indicate substantial heterogeneity, and MR pleiotropy tests did not provide evidence for directional pleiotropy (Supplemental Table S12 & S13), supporting the robustness of this finding. Leave-one-out analyses yielded similar results, suggesting that the association was not driven by any single genetic variant (Supplemental Figure S3-S5).

In reverse-direction two-sample MR with psychiatric conditions as exposures and GD as outcomes, no pairs showed clear causal associations. A borderline association between schizophrenia/other psychotic disorders and uterine fibroids appeared in the primary IVW analysis (Supplemental Table S14) but was not supported by other estimates (Supplemental Table S15). Borderline heterogeneity was observed for anxiety/stress-related disorders and endometriosis (Supplemental Table S16) without evidence of directional pleiotropy (Supplemental Table S17). Leave-one-out analyses gave similar results (Supplemental Figures S6–S8). Results from MR analyses are summarized in Figure 4

**Figure 4:**
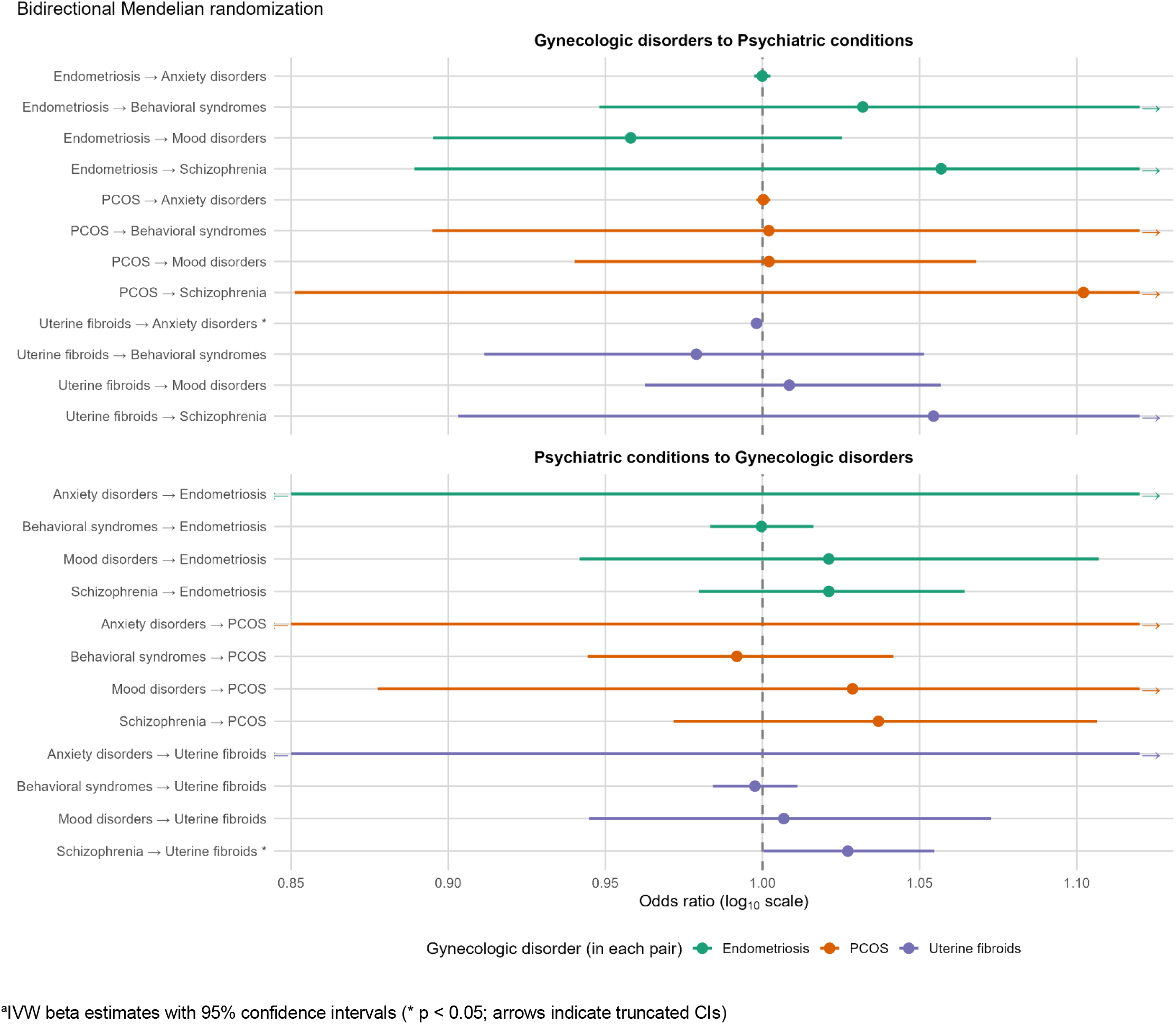
Summary of the two-sample bidirectional Mendelian Randomization analysis^a^. ^a^IVW beta estimates with 95% confidence intervals (* p < 0.05; arrows indicate truncated CIs)

## DISCUSSION

This study is the first to use an integrated causal inference framework combining matched cohort designs, PRS analyses, and bidirectional two-sample MR to characterize observational associations, shared genetic liability, and directionality of the associations between endometriosis, PCOS, fibroids, and the full spectrum of adult-onset psychiatric conditions.

We found that women with uterine fibroids had higher risks of anxiety/stress-related disorders in all three analyses, mood disorders in two analyses, and behavioral syndromes with physical components in the observational analysis. PRS analyses supported associations between genetic liability to fibroids and both mood and anxiety/stress-related disorders, and MR analyses indicated a likely causal effect of fibroid genetic liability on anxiety/stress-related disorders. Women with endometriosis had increased risks of mood disorders, anxiety/stress-related disorders, and behavioral syndromes, whereas women with PCOS had elevated risks of mood and anxiety/stress-related disorders only in the observational analyses. None of the GDs were associated with schizophrenia or other psychotic disorders. These patterns were robust in the sensitivity analyses.

In women with uterine fibroids, we estimated 17–40% higher risks of mood and anxiety/stress- related disorders, underscoring the public health impact of a disorder affecting up to 70-80% of women.^23^ No previous research has examined these risks in this population, limiting direct comparison. Nonetheless, our findings are robust, with affected women having higher risks for multiple psychiatric outcomes. Beyond the observational associations, we identified shared genetic liability between fibroids and both mood and anxiety-/stress-related disorders, with evidence from MR suggesting a likely causal, one-directional effect of fibroids on anxiety-/stress- related disorders (but not the opposite). Mechanistically, molecular biomarkers of uterine fibroids include dysregulated growth factor pathways such as VEGF-A, fibroblast growth factor, IGF-1, and TGF-β. These biomarkers are overexpressed in fibroid tissue and are involved in abnormal tissue repair driven by sex hormones.^24^ Independent psychiatric research implicates overlapping biomarkers—particularly VEGF, FGF2, IGF-1, and TGF-β1—in major depressive disorder and, more variably, anxiety and stress-related conditions, suggesting a shared biological substrate linking fibroids with these psychiatric disorders.^25^

In women with endometriosis, there was an approximately 50% higher risk of mood and anxiety/stress-related disorders. This is consistent with Bliddal et al., who reported a higher incidence of depression among women with endometriosis despite shorter follow-up and limited adjustment for social and economic covariates^26^ and with Koller et al., who showed both phenotypic associations and genetic correlation of endometriosis with depression and anxiety.^3^ The mechanisms that may link endometriosis with depression and anxiety include elevated pro- inflammatory cytokines (e.g., IL-6, TNF-α, and IL-1) in women with endometriosis, which may induce neuroinflammation and alter brain neurotransmitter systems, changing the neurocircuitry and contributing to fatigue, depression, and anxiety.^27,28^

We found that women with PCOS had the highest risks among the GDs studied, with a 50–80% increased risk of mood and anxiety/stress-related disorders. This aligns with the findings of Bliddal et al., and is further supported by meta-analyses and sibling studies reporting elevated rates of depression, anxiety, and bipolar disorder in women with PCOS, although most previous research has focused on premenopausal populations only.^26,29,30^ In PCOS, hyperandrogenism, insulin resistance, and obesity contribute to tissue-specific immune dysregulation across the hypothalamic–pituitary–gonadal axis and key metabolic organs. In these tissues, local inflammatory and oxidative processes—shaped by this androgen excess and metabolic dysfunction—can alter neuroplasticity and stress-system regulation. Together, these interrelated hormonal, metabolic, and immune changes provide a plausible mechanistic basis for the increased vulnerability to depression, anxiety, and stress-related disorders in women with PCOS.^31^

Women with endometriosis or fibroids, but not PCOS, had an elevated risk of behavioral syndromes with physical components. This may be driven by chronic pelvic pain, or hormonal imbalances like estrogen dominance and progesterone resistance, which are prominent in endometriosis and fibroids but less significant in PCOS. These factors can disrupt sleep quality or lead to sexual difficulties.^32,33^ These risks were highest after diagnosis of endometriosis or fibroids, particularly following a fibroids diagnosis. Given the known age-related progression of both conditions, these findings suggest that more advanced stages of endometriosis or fibroids may be associated with a greater risk of behavioral syndromes.

Beyond biological mechanisms, psychosocial factors likely also play a crucial role. Chronic symptoms, including pain and fatigue in endometriosis and fibroids, often lead to emotional distress and frustration, increasing the risk of stress and/or sleep-related conditions.^34^ Social stigma linked to visible symptoms, such as weight gain, hirsutism, and acne in PCOS, can result in low self-esteem and depression.^35^ Delayed diagnoses of GDs and invasive treatments add psychological strain, including prolonged stress and anxiety.^36^ Additionally, the challenges of managing GDs often strain couples’ relationships and sexual intimacy, contributing to loneliness and poorer mental health. Importantly, psychosocial stress feeds back into biology via chronic activation of stress systems, worsening insulin resistance and inflammation, aggravating GD symptoms, and creating a vicious cycle in which psychological distress and biological dysfunction reinforce each other.

### Strengths and Limitations

This study has notable strengths including its innovative methodological approach, integrating matched cohort designs, PRS, and bidirectional two-sample MR to strengthen causal inference and enhance the robustness of findings. PRS analyses capture genetic liability at the individual level, whereas MR provides evidence on the directionality of the association. The long follow-up period and stepwise adjustments further boost the rigor of the analysis, while sensitivity analyses, including conservative definitions of GDs, co-occurrence adjustments, and subgroup analyses, provide consistent and reliable results. However, certain limitations should be considered. The lack of socioeconomic and racial/ethnic diversity limits the generalizability of the findings, particularly as fibroids predominantly affect Black women, while 88% of women with this condition in our sample were White. Additionally, the study does not address the severity or subtypes of GDs, limiting a more nuanced understanding of functional links. While the study accounts for year of birth in its statistical model and matched design, further consideration of temporal shifts in awareness and GDs diagnostic practices in the UK could bring additional insight.

These findings support integrating mental health assessment into gynecologic care. Women with endometriosis, PCOS, or uterine fibroids are at elevated risk of mood and anxiety/stress-related disorders, and those with endometriosis or fibroids are also at increased risk of behavioral syndromes. Routine screening may identify early psychiatric symptoms. A multidisciplinary care model may also be warranted, with gynecologists attentive to psychiatric symptoms and mental health clinicians considering whether gynecologic disease contributes to psychological distress.

Future research should include more diverse populations, with targeted analyses of racial and ethnic subgroups, particularly for uterine fibroids. Studies should examine whether psychiatric risk varies by GD subtype, severity, age at onset, or treatment exposure. Longitudinal studies and intervention trials are needed to clarify pathways and test whether integrated mental health care improves patient-centered outcomes.

## Conclusion

In conclusion, endometriosis, PCOS, and uterine fibroids were associated with increased risks of mood and anxiety- and stress-related disorders in adult women, while endometriosis and fibroids were additionally associated with behavioral syndromes. These associations did not extend to schizophrenia or other psychotic disorders. Among the associations, those involving fibroids appear to be largely driven by shared genetic liability. Both psychosocial and biological mechanisms—chronic pain, hormonal dysregulation, and inflammation—likely underlie these risks. These findings suggest that GDs are closely linked to long-term mental health and should not be managed in isolation. Integrating mental health screening into gynecologic care, alongside more precise subgroup-focused research, may help improve outcomes for affected women.

## Declarations

### Ethics approval

This study involves human participants. The UK Biobank has ethical approval from the NHS National Research Ethics Service (16/NW/0274). All participants provided informed consent prior to taking part in the cohort. Participants gave informed consent to participate in the cohort before taking part.

## Acknowledgements

We thank the participants of the UK Biobank study for their invaluable contribution to this research.

## Author Contributions

KBM and MJ contributed to study conceptualization, design and methodology. KBM were responsible for data management. KBM and ZA did the formal analysis. KBM, ZA and MJ contributed to interpretation. KBM wrote the original draft, and all the authors reviewed and edited revisions. KBM and ZA had access to UKB data. All authors accept responsibility for submission of this study for publication. KBM and MJ are the guarantors of the study. The corresponding author attests that all listed authors meet authorship criteria and that no others meeting the criteria have been omitted.

## Supplementary data

Supplementary data are available in the supplemental document.

## Data availability

Data may be obtained from a third party and are not publicly available. For the UK Biobank study, the data are available on application: https://www.ukbiobank.ac.uk/. The data for the current study were used under license and so are not publicly available.

## Use of artificial intelligence (AI) tools

Claude (Anthropic, PBC) was used to assist with the graphical design and visual formatting of the Evidence Map supplemental Figure 1, and for grammatical checks. The tool was not used in the conceptualization, design, data analysis, or writing this manuscript; Tool: Claude Sonnet 5. Manufacturer: Anthropic, PBC.

