## Supplemental for "Causal Analysis of Gynecologic Disorders and Adult-Onset Psychiatric Conditions: Integrating Matched Cohort, Polygenic Risk Score, and Mendelian Randomization"


**Literature review**

**Supplemental Figure S1.** Evidence map of the associations between female Gynecologic Disorders and adult-onset psychiatric conditions.

**Methods details**

**Supplemental Section S1**. Polygenic Risk Score (PRS) Construction and SNP-based Heritability
**Supplemental Table S1.** Exposure and Related Definitions (Gynecologic Disorders PRS Performance)
**Supplemental Section S2.** GWAS Summary Statistics Processing and Instrument Selection for MR

**Causal framework**

**Supplemental Figure S2**. Causal framework showing the tested associations among gynecologic disorders, psychiatric conditions, and confounders.

**Descriptive Characteristics by Gynecologic Disorder Diagnosis**

**Supplemental Table S2.** Covariates and Psychiatric Conditions by Endometriosis Diagnosis
**Supplemental Table S3.** Covariates and Psychiatric Conditions by PCOS Diagnosis
**Supplemental Table S4.** Covariates and Psychiatric Conditions by Fibroids Diagnosis

**Observational Analyses: Gynecologic Disorders and Psychiatric Outcomes**

**Supplemental Table S5.** Association Between Endometriosis and Psychiatric Outcomes by Time Period
**Supplemental Table S6.** Association Between PCOS and Psychiatric Outcomes by Time Period
**Supplemental Table S7.** Association Between Fibroids and Psychiatric Outcomes by Time Period
**Supplemental Table S8.** Sensitivity Analyses for Associations Between Gynecologic Disorders and Psychiatric Outcomes
**Supplemental Table S9.** PRS-based Definitions of Gynecologic Disorders and Psychiatric Outcomes

**Mendelian Randomization: Gynecologic Disorders → Psychiatric Outcomes**

**Supplemental Table S10.** IVW Two-sample MR (Gynecologic Disorders as Exposures)
**Supplemental Table S11.** MR-Egger, Weighted Median, and Weighted Mode (Gynecologic Disorders as Exposures)
**Supplemental Table S12.** Heterogeneity Statistics (IVW and MR-Egger; Gynecologic Disorders as Exposures)
**Supplemental Table S13**. MR-Egger Intercept Tests for Horizontal Pleiotropy (Gynecologic Disorders as Exposures)
**Supplemental Figure S3.** Scatter Plots: Endometriosis and Psychiatric Outcomes
**Supplemental Figure S4.** Scatter Plots: PCOS and Psychiatric Outcomes
**Supplemental Figure S5.** Scatter Plots: Uterine Fibroids and Psychiatric Outcomes

**Mendelian Randomization: Psychiatric Outcomes → Gynecologic Disorders**

**Supplemental Table S14.** IVW Two-sample MR (Psychiatric Disorders as Exposures)
**Supplemental Table S15.** MR-Egger, Weighted Median, and Weighted Mode (Psychiatric Disorders as Exposures)
**Supplemental Table S16.** Heterogeneity Statistics (IVW and MR-Egger; Psychiatric Disorders as Exposures)
**Supplemental Table S17.** MR-Egger Intercept Tests for Horizontal Pleiotropy (Psychiatric Disorders as Exposures)
**Supplemental Figure S6.** Scatter Plots: Schizophrenia and Gynecologic Disorders
**Supplemental Figure S7.** Scatter Plots: Mood Disorders and Gynecologic Disorders
**Supplemental Figure S8.** Scatter Plots: Anxiety/Stress-related Disorders and Gynecologic Disorders
**Supplemental Figure S9.** Scatter Plots: Behavioral Syndromes and Gynecologic Disorders

**Supplemental Figure S1**. Evidence map of the associations between female Gynecologic Disorders and adult-onset psychiatric conditions.

| Psychiatric Condition Domain | ENDOMETRIOSIS | | | PCOS | | | UTERINE FIBROIDS | | |
|---|---|---|---|---|---|---|---|---|---|
| | Observational (Clinical / Epidemiologic) | Shared Genetics (Genetic Correlation / MR) | Directionality (Temporality / MR) | Observational (Clinical / Epidemiologic) | Shared Genetics (Genetic Correlation / MR) | Directionality (Temporality / MR) | Observational (Clinical / Epidemiologic) | Shared Genetics (Genetic Correlation / MR) | Directionality (Temporality / MR) |
| **Schizophrenia or other psychotic disorders** | **Limited**<br>**Gao 2020** – non-affective psychosis not associated/ Limitations – short follow-up | **Gap**<br>No study identified | **Gap**<br>No study identified | **Limited**<br>**Cesta 2016** – elevated risk in registry/ Limitation- short follow-up | **Conflicting**<br>**Jin 2021** – No association according MR/ Limitations - Genetically predicted PCOS and Schizophrenia<br>**Wen 2025** – MR showing association | **Limited**<br>**Jin 2021** - MR not significant/ / Limitations - Genetically predicted PCOS and Schizophrenia | **Gap**<br>No study identified | **Gap**<br>No study identified | **Gap**<br>No study identified |
| **Mood disorders** (depression, bipolar) | **Evidence**<br>**Chen 2016; Estes 2021; Chen 2020** – depression/bipolar elevated in cohorts | **Evidence**<br>**Adewuyi 2021** – rg with depression, shared loci; **Koller 2023** – shared depression/anxiety liability; **Li 2026** – bipolar correlation | **Evidence**<br>**Adewuyi 2021; Chen 2024; Koller 2023** – bidirectional signal; mood liability→endo in MR | **Limited**<br>**Hung 2014; Berni 2018**– depression, bipolar elevated/ Limitations- very short follow-up | **Conflicting**<br>**Jin 2021** – some null findings; **Zhang 2022** – small PCOS→depression effect; **Li 2026** – shared locus with MDD | **Conflicting**<br>**Hung 2014; Jin 2021** – PCOS precedes mood disorders; MR inconsistent | **Limited**<br>**Shen 2017; Chiuve 2022; Lee 2024** – depression associated/ Limitations- short follow-up | **Limited**<br>**Han 2024; Li 2026** – broad distress MR, not depression-specific (limited) | **Evidence**<br>**Wise 2015** - depression →fibroids **Lee 2024; Han 2024** – bidirectional depression↔fibroid (strongest in fibroids) |
| **Anxiety- and stress-related disorders** | **Evidence**<br>**Chen 2016; Estes 2021** – anxiety elevated | **Evidence**<br>**Koller 2023** – shared genetic liability with anxiety | **Evidence**<br>**Koller 2023** – anxiety liability→endo in MR; | **Limited**<br>**Cooney 2017; Hung 2014** – anxiety elevated/ Limitations- cross-sectional | **Limited**<br>**Jin 2021** – no genetic correlation with anxiety/ Genetically predicted PCOS and anxiety | **Limited**<br>**Jin 2021** – PCOS→anxiety not supported/ Limitations- Genetically predicted PCOS and anxiety | **Limited**<br>**Chiuve 2022;**– anxiety reported/ Limitations- short follow-up | **Gap**<br>Han 2024 No study identified | **Gap**<br>No study identified |
| **Behavioral syndromes with physical components** (eating, sleep) | **Limited**<br>**Gao 2020** – eating disorders associated; sleep disturbance common/ Limitations – short follow-up | **Limited**<br>**Koller 2023** – no genetic support for eating disorders | **Gap**<br>Gap – no study identified | **Limited**<br>**Cesta 2016** – bulimia/eating disorders; / Limitation- short follow-up<br>**Hung 2014** – sleep disorders/ / Limitation- short follow-up | **Limited**<br>**Jin 2021** – shared genetic liability between PCOS and OCD; / Limitations - Genetically predicted PCOS and OCD | **Limited**<br>**Jin 2021** – MR supports PCOS→OCD; / Limitations - Genetically predicted PCOS and OCD | **Gap**<br>No study identified | **Gap**<br>No study identified | **Gap**<br>No study identified |

#### EVIDENCE LEVEL KEY

**Evidence**
Consistent evidence from multiple studies or at least one high quality study

**Limited**
Some evidence but with important limitations, or sparse

**Gap/ conflicting**
No or insufficient PubMed-indexed evidence identified/ / conflicting findings

#### NOTES

- Observational = associations from cohort, case-control, cross-sectional, or registry studies.
- Shared genetics = genetic correlation (rg), GWAS overlap, or polygenic / Mendelian randomization (MR).
- Directionality = temporality from longitudinal studies or causal inference from bidirectional MR.
- Citations are key representative references; full reference list is below.

#### SUMMARY BY DISORDER

**Endometriosis:**
Most complete & consistent evidence across all three layers (observational + genetics + directionality).

**PCOS:**
Strong observational signal but conflicting genetic evidence

**Uterine Fibroids:**
Few evidence; best bidirectional temporality for depression through observational studies, but weak genetic evidence.

**Supplemental section S1. Polygenic Risk Score (PRS) Construction and SNP-based Heritability**

Data processing, clumping and PRS calculations were performed within the UKB Research Analysis Platform. Genotype data quality control was performed using PLINK v2. SNPs with genotype missingness greater than 10%, individuals with genotype missingness greater than 10%, minor allele frequency less than 1%, and those deviating from Hardy-Weinberg equilibrium ($p < 1\times10^{-6}$) were excluded. Individual-level quality control was applied only to participants who met UKB's internal genotyping quality control criteria for principal component analysis. This ensures unrelatedness, concordance between genetic and reported sex, low genotype missingness, and exclusion of heterozygosity outliers, as detailed elsewhere (Bycroft *et al.*, 2018). PRS were calculated using the clumping and thresholding method implemented in PLINK v2.0. GWAS summary statistics for gynecologic disorders (uterine fibroids (Venkatesh *et al.*, 2025), PCOS (Pujol Gualdo *et al.*, 2025) and endometriosis(Kurki *et al.*, 2023)) were obtained from publicly available sources, excluding UKB participants to avoid sample overlap. Clumping was performed with the following parameters: $r^2 < 0.1$ within a 250 kb window, using a p-value threshold of 1 for inclusion. PRS were calculated using these summary statistics across multiple p-value thresholds (e.g., $5\times10^{-8}$, $1\times10^{-5}$, 0.001, 0.01, 0.05, 0.1, 0.5, and 1) with the --score and --q-score-range functions in PLINK. Logistic regression models were then fitted for each PRS threshold, with diagnosis of the corresponding gynecologic disorder as the outcome and the PRS and the first 10 genetic principal components as covariates using R. To evaluate predictive performance, area under the curve (AUC) statistics were calculated for each threshold. The p-value threshold with the highest AUC was selected for each disorder. P-values, AUC, $R^2$, and SNP heritability for each PRS are available in Supplemental Table S1.

<u>SNP-based heritability</u>

To estimate SNP-based heritability (SNP-$h^2$) for the gynecologic disorders_, we used the linkage disequilibrium score regression (LDSC) method. Prior to the analysis, we converted the files to LDSC format (implemented in munge_sumstats.py). The LD scores were pre-computed based on 1000 Genomes Project Phase 3 reference data on individuals of European ancestry.

**Table S1: Exposure and related definitions**

| **Gynecologic disorders** | **ICD-10 codes** | **Polygenic Risk Score** | | | |
|---|---|---|---|---|---|
| | | **P-value** | **AUC** | **Nagelkerke R2** | **SNP Heritability** |
| Endometriosis | N80 | 0.0001 | 0.553 | 0.002 | 0.0912 |
| Polycystic ovary syndrome | E28.2 | 0.001 | 0.582 | 0.002 | 0.015 |
| Uterine fibroids | D25 Leiomyoma of uterus | 0.5 | 0.730 | 0.156 | 0.0221 |

**Supplemental Figure S2. Causal framework showing the tested associations among Gynecologic disorders, psychiatric conditions, and confounders.**

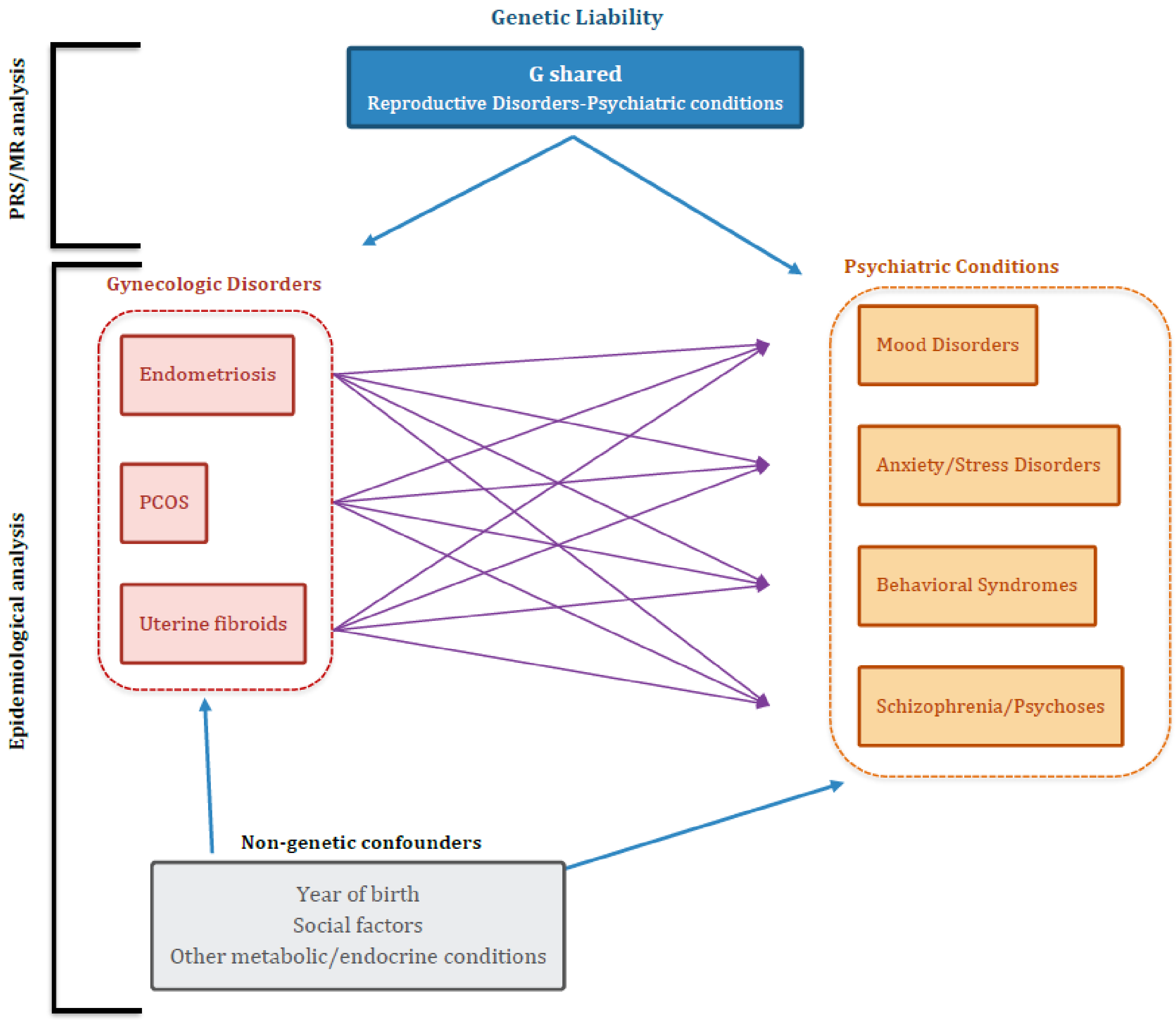

**Supplemental section S2. GWAS Summary Statistics Processing and Instrument Selection for MR**

We used harmonized genome-wide association summary statistics for each gynecologic disorder (statistics for endometriosis, uterine fibroids and PCOS)(Kurki *et al.*, 2023; Pujol Gualdo *et al.*, 2025; Venkatesh *et al.*, 2025). To ensure the independence of the variants, we clumped the identified SNPs using the 1000 Genomes Project reference panel for European populations. We applied a clumping window of 10,000 kilobases and an LD r2 cutoff of 0.001. For primary analyses (gynecologic disorders as exposures), SNPs were selected at genome-wide significance ($p < 5 \times 10^{-8}$). For reverse-direction analyses (psychiatric disorders as exposures), a more liberal threshold ($p < 5 \times 10^{-6}$) was applied to increase the number of available instruments. Within each LD window, the SNP with the lowest p-value was retained. During threshold. Additionally, effect alleles were aligned between datasets, excluding SNPs with ambiguous allele pairings and removing palindromic SNPs with intermediate allele frequencies to prevent strand errors.

**Supplemental Table S2: Covariates and psychiatric conditions according to endometriosis diagnosis status.**

| Variables | With endometriosis diagnosis | | Without endometriosis diagnosis | |
|---|---|---|---|---|
| | N=10755 | % | N=10755 | % |
| **Index of Multiple Deprivation quintile[a]** | | | | |
| 1st quintile (richest) | 1909 | 17.8 | 1908 | 17.8 |
| 2nd quintile | 2057 | 19.2 | 2057 | 19.2 |
| 3rd quintile | 2249 | 21 | 2249 | 21 |
| 4th quintile | 2242 | 20.9 | 2242 | 20.9 |
| 5th quintile (poorest) | 2298 | 21.4 | 2299 | 21.4 |
| **Income** | | | | |
| Less than 18,000 | 2042 | 19 | 1979 | 18.5 |
| 18,000 to 30,999 | 2238 | 20.9 | 2236 | 20.8 |
| 31,000 to 51,999 | 2444 | 22.8 | 2425 | 22.6 |
| 52,000 to 100,000 | 1898 | 17.7 | 1872 | 17.5 |
| Greater than 100,000 | 374 | 3.5 | 508 | 4.8 |
| Prefer not to answer | 1189 | 11.1 | 1087 | 10.2 |
| Missing value | 570 | 5.3 | 648 | 6.1 |
| **Degree** | | | | |
| A levels/AS levels or equivalent | 305 | 2.9 | 328 | 3.1 |
| College or University degree | 1094 | 10.2 | 1169 | 10.9 |
| CSEs or equivalent | 1116 | 10.4 | 960 | 9 |
| None of the above | 1428 | 13.3 | 1422 | 13.3 |
| NVQ or HND or HNC or equivalent | 1586 | 14.8 | 1477 | 13.8 |
| O levels/GCSEs or equivalent | 2020 | 18.8 | 2065 | 19.3 |
| Other professional qualifications (nursing, teaching) | 3036 | 28.3 | 3129 | 29.1 |
| Missing value | 170 | 1.6 | 205 | 2 |
| **Race** | | | | |
| White | 9775 | 90.9 | 9590 | 89.2 |
| Asian or Asian British | 383 | 3.6 | 515 | 4.8 |
| Black or Black British | 95 | 0.9 | 74 | 0.7 |
| More than one race | 342 | 3.2 | 394 | 3.7 |
| Other ethnic groups | 107 | 1 | 118 | 1.1 |
| Missing value | 53 | 0.5 | 64 | 0.6 |
| **Migration status** | | | | |
| No | 9894 | 92 | 9696 | 90.2 |
| Yes | 847 | 7.9 | 1035 | 9.7 |
| Missing value | 14 | 0.2 | 24 | 0.3 |
| **Metabolic/endocrine conditions** | | | | |
| No | 6615 | 61.6 | 7311 | 68 |
| Yes | 4140 | 38.5 | 3444 | 32.1 |
| **Diagnosis of PCOS** | | | | |
| No | 10386 | 96.6 | 10629 | 98.9 |
| Yes | 369 | 3.5 | 126 | 1.2 |
| **Diagnosis of Fibroids** | | | | |
| No | 7182 | 66.8 | 9974 | 92.8 |
| Yes | 3573 | 33.3 | 781 | 7.3 |
| **Schizophrenia and other psychotic disorders (F20-F29) any time[b]** | | | | |

| | | | | |
|---|---|---|---|---|
| No | 10698 | 99.5 | 10704 | 99.6 |
| Yes | 57 | 0.6 | 51 | 0.5 |
| **Mood disorders (F30-F39) any time[b]** | | | | |
| No | 8301 | 77.2 | 8976 | 83.5 |
| Yes | 2454 | 22.9 | 1779 | 16.6 |
| **Anxiety- or stress-related disorders (F40-F48) any time[b]** | | | | |
| No | 8487 | 79 | 9110 | 84.8 |
| Yes | 2268 | 21.1 | 1645 | 15.3 |
| **Behavioral syndromes with physical components (F50-F59) any time[b]** | | | | |
| No | 10547 | 98.1 | 10584 | 98.5 |
| Yes | 208 | 2 | 171 | 1.6 |
| **Primary care linkage[a]** | | | | |
| No | 4784 | 44.5 | 4784 | 44.5 |
| Yes | 5971 | 55.6 | 5971 | 55.6 |
| **Numeric variables mean (±SD, median; Q1-Q3)** | | | | |
| Age at recruitment | 53.2 ± 8, 53; 47-60 | | 53.2 ± 8, 52; 46-60 | |
| Year of birth[a] | 1955 ± 8, 1956; 1948-1962 | | 1955 ± 8, 1956; 1948-1962 | |
| Age at the first diagnosis of endometriosis | 43.0 ±11.4, 42.6; 35.3-49.8 | | X | |
| Age at first diagnosis of schizophrenia and other psychotic disorders | 53.2 ± 14.6, 53.8; 42.9-64 | | 53 ± 13.3, 51.9; 44-62.9 | |
| Age at first diagnosis of mood disorders | 45.5 ± 14.5, 46.3; 36.2-55 | | 46.4 ± 14.7, 47.2; 36.6-56.6 | |
| Age at first diagnosis of anxiety- or stress-related disorders | 50.9 ± 13, 51.1; 43.2-59.3 | | 51.4 ± 13.5, 51.7; 42.6-60.3 | |
| Age at first diagnosis of behavioral syndromes with physical components | 38.4 ± 12, 37.9; 30.1-47.4 | | 35.7 ± 13.5, 33.5; 26.5-44.6 | |

[a]Matching variables

[b]Unrestricted follow-up, encompassing the entire duration of follow-up available in the UK Biobank

**Supplemental Table S3: Covariates and psychiatric conditions according to PCOS diagnosis status.**

| Variables | With PCOS diagnosis | | Without PCOS diagnosis | |
|---|---|---|---|---|
| | N=2536 | % | N=12680 | % |
| **Index of Multiple Deprivation quintile[a]** | | | | |
| 1st quintile (richest) | 388 | 15.3 | 1940 | 15.3 |
| 2nd quintile | 469 | 18.5 | 2345 | 18.5 |
| 3rd quintile | 550 | 21.7 | 2750 | 21.7 |
| 4th quintile | 566 | 22.4 | 2830 | 22.4 |
| 5th quintile (poorest) | 563 | 22.3 | 2815 | 22.3 |
| **Income** | | | | |
| Less than 18,000 | 505 | 20 | 2243 | 17.7 |
| 18,000 to 30,999 | 481 | 19 | 2540 | 20.1 |
| 31,000 to 51,999 | 607 | 24 | 2984 | 23.6 |
| 52,000 to 100,000 | 467 | 18.5 | 2399 | 19 |
| Greater than 100,000 | 125 | 5 | 602 | 4.8 |
| Prefer not to answer | 231 | 9.2 | 1245 | 9.9 |
| Missing value | 120 | 4.8 | 667 | 5.3 |
| **Degree** | | | | |
| A levels/AS levels or equivalent | 86 | 3.4 | 400 | 3.2 |
| College or University degree | 300 | 11.9 | 1405 | 11.1 |
| CSEs or equivalent | 245 | 9.7 | 1202 | 9.5 |
| None of the above | 274 | 10.9 | 1588 | 12.6 |
| NVQ or HND or HNC or equivalent | 332 | 13.1 | 1832 | 14.5 |
| O levels/GCSEs or equivalent | 479 | 18.9 | 2418 | 19.1 |
| Other professional qualifications (nursing, teaching) | 780 | 30.8 | 3693 | 29.2 |
| Missing value | 40 | 1.6 | 142 | 1.2 |
| **Race** | | | | |
| White | 2218 | 87.5 | 11352 | 89.6 |
| Asian or Asian British | 126 | 5 | 594 | 4.7 |
| Black or Black British | 34 | 1.4 | 81 | 0.7 |
| More than one race | 105 | 4.2 | 477 | 3.8 |
| Other ethnic groups | 35 | 1.4 | 112 | 0.9 |
| Missing value | 18 | 0.8 | 64 | 0.6 |
| **Migration status** | | | | |
| No | 2246 | 88.6 | 11554 | 91.2 |
| Yes | 284 | 11.2 | 1108 | 8.8 |
| Missing value | 6 | 0.3 | 18 | 0.2 |
| **Metabolic/endocrine conditions** | | | | |
| No | 1382 | 54.5 | 8754 | 69.1 |
| Yes | 1154 | 45.6 | 3926 | 31.0 |
| **Diagnosis of endometriosis** | | | | |
| No | 2167 | 85.4 | 11997 | 94.6 |
| Yes | 369 | 14.5 | 683 | 5.4 |
| **Diagnosis of Fibroids** | | | | |
| No | 1816 | 71.7 | 11029 | 87 |
| Yes | 720 | 28.4 | 1651 | 13.1 |
| **Schizophrenia and other psychotic disorders (F20-F29) any time[b]** | | | | |

| | | | | |
|---|---|---|---|---|
| No | 2516 | 99.3 | 12614 | 99.5 |
| Yes | 20 | 0.8 | 66 | 0.6 |
| **Mood disorders (F30-F39) any time[b]** | | | | |
| No | 1859 | 73.4 | 10202 | 80.5 |
| Yes | 677 | 26.7 | 2478 | 19.6 |
| **Anxiety- or stress-related disorders (F40-F48) any time[b]** | | | | |
| No | 1777 | 70.1 | 10194 | 80.4 |
| Yes | 759 | 30 | 2486 | 19.7 |
| **Behavioral syndromes with physical components (F50-F59) any time[b]** | | | | |
| No | 2454 | 96.8 | 12367 | 97.6 |
| Yes | 82 | 3.3 | 313 | 2.5 |
| **Primary care linkage[a]** | | | | |
| No | 269 | 10.7 | 1345 | 10.7 |
| Yes | 2267 | 89.4 | 11335 | 89.4 |
| **Numeric variables mean (±SD, median; Q1-Q3)** | | | | |
| Age at recruitment | 51.7 ± 8.3, 50; 45-59 | | 51.6 ± 8.2, 50; 44-58 | |
| Year of birth[a] | 1956.7 ± 8.3, 1958; 1950-1964 | | 1956.7 ± 8.3, 1958; 1950-1964 | |
| Age at the first diagnosis of PCOS | 44.8 ± 13.0, 45; 35-53 | | X | |
| Age at first diagnosis of schizophrenia and other psychotic disorders | 49.6 ± 15.2, 49.2; 35.7-59.2 | | 48.6 ± 14.2, 49.2; 41-57.7 | |
| Age at first diagnosis of mood disorders | 44.5 ± 14.1, 45.5; 35.1-53.7 | | 44.5 ± 13.8, 44.8; 35.6-53.9 | |
| Age at first diagnosis of anxiety- or stress-related disorders | 47.7 ± 12.7, 48.2; 39.9-56.1 | | 48.9 ± 12.5, 48.7; 40.9-57.1 | |
| Age at first diagnosis of behavioral syndromes with physical components | 36.3 ± 12.7, 36; 27.2-46.1 | | 35.3 ± 12.7, 33.5; 26.3-43 | |

[a]Matching variables

[b]Unrestricted follow-up, encompassing the entire duration of follow-up available in the UK Biobank

**Supplemental Table S4: Covariates and psychiatric conditions according to fibroids diagnosis status.**

| Variables | With Fibroids diagnosis | | Without Fibroids diagnosis | |
|---|---|---|---|---|
| | N=26057 | % | N=26057 | % |
| **Index of Multiple Deprivation quintile[a]** | | | | |
| 1st quintile (richest) | 4507 | 17.3 | 4506 | 17.3 |
| 2nd quintile | 4816 | 18.5 | 4816 | 18.5 |
| 3rd quintile | 5463 | 21 | 5463 | 21 |
| 4th quintile | 5734 | 22.1 | 5735 | 22.1 |
| 5th quintile (poorest) | 5537 | 21.3 | 5537 | 21.3 |
| **Income** | | | | |
| Less than 18,000 | 4957 | 19.1 | 4948 | 19 |
| 18,000 to 30,999 | 5591 | 21.5 | 5550 | 21.3 |
| 31,000 to 51,999 | 5926 | 22.8 | 5788 | 22.3 |
| 52,000 to 100,000 | 4250 | 16.4 | 4300 | 16.6 |
| Greater than 100,000 | 794 | 3.1 | 1078 | 4.2 |
| Prefer not to answer | 2942 | 11.3 | 2747 | 10.6 |
| Missing value | 1597 | 6.2 | 1646 | 6.4 |
| **Degree** | | | | |
| A levels/AS levels or equivalent | 732 | 2.9 | 728 | 2.8 |
| College or University degree | 2843 | 11 | 2810 | 10.8 |
| CSEs or equivalent | 2295 | 8.9 | 2211 | 8.5 |
| None of the above | 3664 | 14.1 | 3814 | 14.7 |
| NVQ or HND or HNC or equivalent | 3433 | 13.2 | 3389 | 13.1 |
| O levels/GCSEs or equivalent | 4902 | 18.9 | 5137 | 19.8 |
| Other professional qualifications (nursing, teaching) | 7719 | 29.7 | 7514 | 28.9 |
| Missing value | 469 | 1.8 | 454 | 1.8 |
| **Race** | | | | |
| White | 22989 | 88.3 | 23421 | 89.9 |
| Asian or Asian British | 1162 | 4.5 | 1162 | 4.5 |
| Black or Black British | 209 | 0.9 | 144 | 0.6 |
| More than one race | 1118 | 4.3 | 903 | 3.5 |
| Other ethnic groups | 412 | 1.6 | 274 | 1.1 |
| Missing value | 167 | 0.7 | 153 | 0.6 |
| **Migration status** | | | | |
| No | 22941 | 88.1 | 23807 | 91.4 |
| Yes | 3075 | 11.9 | 2202 | 8.5 |
| Missing value | 41 | 0.2 | 48 | 0.2 |
| **Metabolic/endocrine conditions** | | | | |
| No | 15760 | 60.5 | 17450 | 67 |
| Yes | 10297 | 39.6 | 8607 | 33.1 |
| **Diagnosis of endometriosis** | | | | |
| No | 22484 | 86.3 | 25192 | 96.7 |
| Yes | 3573 | 13.7 | 865 | 3.3 |
| **Diagnosis of PCOS** | | | | |
| No | 25337 | 97.3 | 25785 | 99 |
| Yes | 720 | 2.8 | 272 | 1.1 |
| **Schizophrenia and other psychotic disorders (F20-F29) any time[b]** | | | | |

| | | | | |
|---|---|---|---|---|
| No | 25922 | 99.5 | 25912 | 99.5 |
| Yes | 135 | 0.6 | 145 | 0.6 |
| **Mood disorders (F30-F39) any time[b]** | | | | |
| No | 21018 | 80.7 | 21626 | 83 |
| Yes | 5039 | 19.4 | 4431 | 17.1 |
| **Anxiety- or stress-related disorders (F40-F48) any time[b]** | | | | |
| No | 20605 | 79.1 | 21891 | 84.1 |
| Yes | 5452 | 21 | 4166 | 16 |
| **Behavioral syndromes with physical components (F50-F59) any time[b]** | | | | |
| No | 25515 | 98 | 25612 | 98.3 |
| Yes | 542 | 2.1 | 445 | 1.8 |
| **Primary care linkage[a]** | | | | |
| No | No | 9364 | 36 | 9364 |
| Yes | Yes | 16693 | 64.1 | 16693 |
| **Numeric variables mean (±SD, median; Q1-Q3)** | | | | |
| Age at recruitment | 54.5 ± 7.9, 54; 48-61 | | 54.4 ± 7.9, 54; 48-61 | |
| Year of birth[a] | 1953.8 ± 7.9, 1954; 1947-1960 | | 1953.8 ± 7.9, 1954; 1947-1960 | |
| Age at the first diagnosis of fibroids | 50.8 ± 8.9, 50; 45-55 | | x | |
| Age at first diagnosis of schizophrenia and other psychotic disorders | 54.6 ± 13.1, 56.5; 48.9-64.6 | | 51.8 ± 16.1, 53.2; 41.5-62.7 | |
| Age at first diagnosis of mood disorders | 47.1 ± 14.7, 48.2; 37.3-57.1 | | 46.7 ± 15.1, 47.8; 36.6-57.1 | |
| Age at first diagnosis of anxiety- or stress-related disorders | 51.4 ± 13.2, 51.8; 43.5-60.2 | | 51.6 ± 13.8, 52.6; 43.5-60.7 | |
| Age at first diagnosis of behavioral syndromes with physical components | 40.8 ± 14.4, 40.5; 29-52.9 | | 36.9 ± 14.3, 33.6; 26-46.7 | |

[a]Matching variables

[b]Unrestricted follow-up, encompassing the entire duration of follow-up available in the UK Biobank

### Supplemental Table S5: Association Between Endometriosis and Psychiatric Outcomes Through Stepwise Adjustment by Time Period

| Psychiatric Outcomes | Time Period | Model | Odds Ratio (OR) | 95% Confidence Interval (CI) | |
|---|---|---|---|---|---|
| **Schizophrenia & other psychotic disorders** | Any Time | Baseline[a] | 1.12 | 0.77 | 1.63 |
| | | Adjusted for SES[b] | 1.07 | 0.51 | 2.23 |
| | | Fully Adjusted[c] | 1.05 | 0.5 | 2.21 |
| | After | Baseline[a] | 0.98 | 0.63 | 1.52 |
| | | Adjusted for SES[b] | 0.66 | 0.25 | 1.72 |
| | | Fully Adjusted[c] | 0.66 | 0.25 | 1.72 |
| **Mood disorders** | Any Time | Baseline[a] | 1.51 | 1.41 | 1.62 |
| | | Adjusted for SES[b] | 1.51 | 1.4 | 1.63 |
| | | Fully Adjusted[c] | 1.51 | 1.4 | 1.63 |
| | After | Baseline[a] | 1.37 | 1.26 | 1.49 |
| | | Adjusted for SES[b] | 1.38 | 1.26 | 1.52 |
| | | Fully Adjusted[c] | 1.38 | 1.26 | 1.52 |
| **Anxiety- and stress-related disorders** | Any Time | Baseline[a] | 1.51 | 1.4 | 1.62 |
| | | Adjusted for SES[b] | 1.5 | 1.38 | 1.62 |
| | | Fully Adjusted[c] | 1.49 | 1.38 | 1.62 |
| | After | Baseline[a] | 1.42 | 1.31 | 1.54 |
| | | Adjusted for SES[b] | 1.45 | 1.33 | 1.59 |
| | | Fully Adjusted[c] | 1.45 | 1.32 | 1.59 |
| **Behavioral syndromes with physical components** | Any Time | Baseline[a] | 1.23 | 1 | 1.51 |
| | | Adjusted for SES[b] | 1.28 | 1.01 | 1.62 |
| | | Fully Adjusted[c] | 1.28 | 1.01 | 1.62 |
| | After | Baseline[a] | 1.66 | 1.2 | 2.29 |
| | | Adjusted for SES[b] | 2.05 | 1.37 | 3.08 |
| | | Fully Adjusted[c] | 2.05 | 1.37 | 3.08 |

[a]Multivariable conditional logistic regression adjusted for primary care data status, IMD, and birth year.

[b]Multivariable conditional logistic regression adjusted for primary care data status, IMD, and birth year, race, education, household income, migration status.

[c]Multivariable conditional logistic regression adjusted for primary care data status, IMD, and birth year, race, education, household income, migration status, metabolic/endocrine conditions status (obesity, diabetes, thyroid disorders, and other metabolic disorders).

### Supplemental Table S6: Association Between PCOS and Psychiatric Outcomes Through Stepwise Adjustment by Time Period

| Psychiatric Outcome | Time Period | Model | Odds Ratio (OR) | 95% Confidence Interval (CI) | |
|---|---|---|---|---|---|
| **Schizophrenia & other psychotic disorders** | Any Time | Baseline[a] | 1.52 | 0.92 | 2.52 |
| | | Adjusted for SES[b] | 1.54 | 0.82 | 2.87 |
| | | Fully Adjusted[c] | 1.46 | 0.78 | 2.74 |
| | After | Baseline | 1.49 | 0.76 | 2.94 |
| | | Adjusted for SES | 1.23 | 0.49 | 3.08 |
| | | Fully Adjusted | 1.15 | 0.45 | 2.95 |
| **Mood disorders** | Any Time | Baseline | 1.51 | 1.37 | 1.67 |
| | | Adjusted for SES | 1.51 | 1.35 | 1.67 |
| | | Fully Adjusted | 1.47 | 1.32 | 1.64 |
| | After | Baseline | 1.43 | 1.25 | 1.63 |
| | | Adjusted for SES | 1.43 | 1.25 | 1.65 |
| | | Fully Adjusted | 1.4 | 1.22 | 1.61 |
| **Anxiety- and stress-related disorders** | Any Time | Baseline | 1.78 | 1.61 | 1.96 |
| | | Adjusted for SES | 1.81 | 1.63 | 2.01 |
| | | Fully Adjusted | 1.79 | 1.62 | 1.99 |
| | After | Baseline | 1.5 | 1.33 | 1.7 |
| | | Adjusted for SES | 1.51 | 1.34 | 1.72 |
| | | Fully Adjusted | 1.51 | 1.33 | 1.71 |
| **Behavioral syndromes with physical components** | Any Time | Baseline | 1.33 | 1.04 | 1.71 |
| | | Adjusted for SES | 1.3 | 1.00 | 1.7 |
| | | Fully Adjusted | 1.31 | 1.00 | 1.7 |
| | After | Baseline | 1.23 | 0.8 | 1.89 |
| | | Adjusted for SES | 1.13 | 0.7 | 1.81 |
| | | Fully Adjusted | 1.13 | 0.7 | 1.81 |

[a]Multivariable conditional logistic regression adjusted for primary care data status, IMD, and birth year.
[b]Multivariable conditional logistic regression adjusted for primary care data status, IMD, and birth year, race, education, household income, migration status.
[c]Multivariable conditional logistic regression adjusted for primary care data status, IMD, and birth year, race, education, household income, migration status, metabolic/endocrine conditions status (obesity, diabetes, thyroid disorders, and other metabolic disorders).

**Supplemental Table S7: Association Between Fibroids and Psychiatric Outcomes Through Stepwise Adjustment by Time Period.**

| Psychiatric Outcome | Time Period | Model | Odds Ratio (OR) | 95% Confidence Interval (CI) | |
|---|---|---|---|---|---|
| **Schizophrenia & other psychotic disorders** | Any Time | Baseline[a] | 0.93 | 0.74 | 1.18 |
| | | Adjusted for SES[b] | 0.81 | 0.58 | 1.15 |
| | | Fully Adjusted[c] | 0.8 | 0.57 | 1.14 |
| | After | Baseline | 1.04 | 0.77 | 1.4 |
| | | Adjusted for SES | 0.93 | 0.57 | 1.53 |
| | | Fully Adjusted | 0.91 | 0.55 | 1.5 |
| **Mood disorders** | Any Time | Baseline | 1.17 | 1.12 | 1.23 |
| | | Adjusted for SES | 1.18 | 1.12 | 1.24 |
| | | Fully Adjusted | 1.17 | 1.11 | 1.23 |
| | After | Baseline | 1.1 | 1.03 | 1.17 |
| | | Adjusted for SES | 1.11 | 1.04 | 1.2 |
| | | Fully Adjusted | 1.1 | 1.03 | 1.19 |
| **Anxiety- and stress-related disorders** | Any Time | Baseline | 1.41 | 1.34 | 1.47 |
| | | Adjusted for SES | 1.41 | 1.34 | 1.49 |
| | | Fully Adjusted | 1.4 | 1.34 | 1.48 |
| | After | Baseline | 1.3 | 1.23 | 1.38 |
| | | Adjusted for SES | 1.32 | 1.23 | 1.4 |
| | | Fully Adjusted | 1.31 | 1.23 | 1.4 |
| **Behavioral syndromes with physical components** | Any Time | Baseline | 1.22 | 1.08 | 1.39 |
| | | Adjusted for SES | 1.29 | 1.12 | 1.49 |
| | | Fully Adjusted | 1.29 | 1.12 | 1.48 |
| | After | Baseline | 2.01 | 1.57 | 2.57 |
| | | Adjusted for SES | 2.27 | 1.68 | 3.06 |
| | | Fully Adjusted | 2.25 | 1.67 | 3.03 |

[a]Multivariable conditional logistic regression adjusted for primary care data status, IMD, and birth year.

[b]Multivariable conditional logistic regression adjusted for primary care data status, IMD, and birth year, race, education, household income, migration status.

[c]Multivariable conditional logistic regression adjusted for primary care data status, IMD, and birth year, race, education, household income, migration status, metabolic/endocrine conditions status (obesity, diabetes, thyroid disorders, and other metabolic disorders).

**Supplemental Table S8: Sensitivity analysis of the Association Between Gynecologic disorder and Psychiatric Outcomes in the overall study Period.**

| Psychiatric Outcome | Gynecologic disorder | Analysis Type | | | |
|---|---|---|---|---|---|
| | | Main Analysis[a] | Adjusted for Co-occurrence of RD[b,] | Conservative definition of RD[c] | Restricted to women with Primary care data[d] |
| **Schizophrenia & other psychotic disorders** | Endometriosis | 1.05 [0.5; 2.21] | 0.62 [0.25; 1.51] | 0.99 [0.65; 1.52] | 1.01 [0.57; 1.82] |
| | Fibroids | 0.8 [0.57; 1.14] | 0.84 [0.58; 1.19] | 0.91 [0.68; 1.22] | 0.64 [0.4; 1.02] |
| | PCOS | 1.46 [0.78; 2.74] | 1.51 [0.8; 2.85] | 1.8 [1; 3.24] | 1.64 [0.86; 3.14] |
| **Mood disorders** | Endometriosis | 1.51 [1.4; 1.63] | 1.5 [1.38; 1.62] | 1.52 [1.42; 1.63] | 1.49 [1.36; 1.63] |
| | Fibroids | 1.17 [1.11; 1.23] | 1.16 [1.1; 1.22] | 1.16 [1.11; 1.21] | 1.17 [1.1; 1.24] |
| | PCOS | 1.47 [1.32; 1.64] | 1.43 [1.29; 1.6] | 1.49 [1.32; 1.69] | 1.43 [1.28; 1.6] |
| **Anxiety- and stress-related disorders** | Endometriosis | 1.49 [1.38; 1.62] | 1.45 [1.33; 1.57] | 1.38 [1.29; 1.48] | 1.47 [1.35; 1.61] |
| | Fibroids | 1.4 [1.34; 1.48] | 1.4 [1.33; 1.47] | 1.37 [1.31; 1.44] | 1.43 [1.35; 1.51] |
| | PCOS | 1.79 [1.62; 1.99] | 1.77 [1.59; 1.96] | 1.7 [1.51; 1.92] | 1.79 [1.61; 1.99] |
| **Behavioral syndromes with physical components** | Endometriosis | 1.28 [1.01; 1.62] | 1.3 [1.02; 1.66] | 1.25 [1.02; 1.53] | 1.29 [1.02; 1.64] |
| | Fibroids | 1.29 [1.12; 1.48] | 1.29 [1.12; 1.48] | 1.26 [1.1; 1.43] | 1.31 [1.13; 1.52] |
| | PCOS | 1.31 [1; 1.7] | 1.28 [0.98; 1.68] | 1.26 [0.91; 1.74] | 1.28 [0.98; 1.68] |

[a]Multivariable conditional logistic regression adjusted for primary care data status, IMD, and birth year, race, education level, household income, migration status, metabolic/endocrine conditions status (obesity, diabetes, thyroid disorders, and other metabolic disorders). Diagnosed vs non-diagnosed respectively Endometriosis (n 10755 vs 10755), PCOS (n 2536 vs 12680) Fibroids (n 26057 vs 26057).

[b]The main analysis model was additionally adjusted for Fibroids and PCOS, respectively, when estimating the association of Endometriosis with outcomes, for Fibroids and Endometriosis when assessing PCOS, and for PCOS and Endometriosis when evaluating Fibroids. Diagnosed vs non-diagnosed respectively Endometriosis (n 10755 vs 10755), PCOS (n 2536 vs 12680) Fibroids (n 26057 vs 26057)

[c]Main analysis model in a matched cohort with conservative definition of diagnosed vs non-diagnosed respectively endometriosis (n 6981 vs 41886), PCOS (n 1615 vs19380) and fibroids (n 21932 vs 43864).

[d] Diagnosed vs non-diagnosed respectively Endometriosis (n 5971 vs 5971), PCOS (n 2267 vs 11335) and Fibroids (n 16693 vs 16693)

**Supplemental Table S9: Analysis of the Association Between Gynecologic disorder and Psychiatric Outcomes in the overall study Period using PRS based definition of Gynecologic disorders.**

| Psychiatric Outcome | Gynecologic disorder | Odds Ratio (OR) | 95% Confidence Interval (CI) | |
|---|---|---|---|---|
| **Schizophrenia & other psychotic disorders** | **Endometriosis** | 1.01 | 0.95 | 1.08 |
| | **Fibroids** | 1.01 | 0.95 | 1.07 |
| | **PCOS** | 1.04 | 0.98 | 1.11 |
| **Mood disorders** | **Endometriosis** | 1.02 | 1.00 | 1.03 |
| | **Fibroids** | 1.03 | 1.02 | 1.05 |
| | **PCOS** | 1.02 | 1.00 | 1.03 |
| **Anxiety- and stress-related disorders** | **Endometriosis** | 1.00 | 0.99 | 1.01 |
| | **Fibroids** | 1.04 | 1.02 | 1.05 |
| | **PCOS** | 1.01 | 0.99 | 1.02 |
| **Behavioral syndromes with physical components** | **Endometriosis** | 1.01 | 0.98 | 1.05 |
| | **Fibroids** | 1.00 | 0.96 | 1.04 |
| | **PCOS** | 0.98 | 0.94 | 1.01 |

**Supplemental Table S10: Two-sample Mendelian Randomization analysis using inverse variance weighting testing gynecologic disorders as exposures and psychiatric disorders as outcomes.**

| Gynecologic disorder | Psychiatric Outcome | B | SE | p-value |
|---|---|---|---|---|
| Endometriosis | Schizophrenia & other psychotic disorders | 0.055 | 0.088 | 0.530 |
| | Mood disorders | -0.043 | 0.035 | 0.216 |
| | Anxiety- and stress-related disorders | 0.000 | 0.001 | 0.971 |
| | Behavioral syndromes with physical components | 0.031 | 0.043 | 0.467 |
| PCOS | Schizophrenia & other psychotic disorders | 0.097 | 0.132 | 0.461 |
| | Mood disorders | 0.002 | 0.033 | 0.949 |
| | Anxiety- and stress-related disorders | 0.000 | 0.001 | 0.813 |
| | Behavioral syndromes with physical components | 0.002 | 0.058 | 0.972 |
| Fibroids | Schizophrenia & other psychotic disorders | 0.053 | 0.079 | 0.502 |
| | Mood disorders | 0.008 | 0.024 | 0.721 |
| | Anxiety- and stress-related disorders | -0.002 | 0.001 | 0.016 |
| | Behavioral syndromes with physical components | -0.021 | 0.036 | 0.559 |

B is the inverse-variance weighted two-sample Mendelian randomization effect estimate for the association between genetically predicted gynecologic disorders and psychiatric outcomes, expressed on the original GWAS scales of the exposure and outcome.

SE = standard error.

**Supplemental Table S11: Two-sample Mendelian Randomization analyses (MR-Egger, weighted median, and weighted mode) estimating the effect of genetic liability to gynecologic disorders on psychiatric outcomes.**

| Gynecologic disorder | Psychiatric Outcome | MR-Egger | | | Weighted median | | | Weighted mode | | |
|---|---|---|---|---|---|---|---|---|---|---|
| | | B | SE | p-value | B | SE | p-value | B | SE | p-value |
| Endometriosis | Schizophrenia & other psychotic disorders | -0.181 | 0.249 | 0.474 | -0.134 | 0.129 | 0.300 | -0.238 | 0.180 | 0.197 |
| | Mood disorders | 0.041 | 0.095 | 0.673 | -0.039 | 0.044 | 0.373 | -0.024 | 0.059 | 0.680 |
| | Anxiety- and stress-related disorders | 0.002 | 0.004 | 0.704 | 0.002 | 0.002 | 0.255 | 0.003 | 0.002 | 0.217 |
| | Behavioral syndromes with physical components | 0.105 | 0.122 | 0.396 | 0.099 | 0.066 | 0.131 | 0.114 | 0.097 | 0.250 |
| PCOS | Schizophrenia & other psychotic disorders | -0.018 | 0.364 | 0.963 | 0.093 | 0.156 | 0.550 | -0.017 | 0.211 | 0.938 |
| | Mood disorders | -0.018 | 0.091 | 0.850 | 0.023 | 0.040 | 0.571 | 0.028 | 0.051 | 0.598 |
| | Anxiety- and stress-related disorders | -0.001 | 0.004 | 0.879 | 0.000 | 0.002 | 0.936 | 0.000 | 0.002 | 0.928 |
| | Behavioral syndromes with physical components | -0.154 | 0.144 | 0.327 | -0.076 | 0.070 | 0.277 | -0.085 | 0.088 | 0.368 |
| Fibroids | Schizophrenia & other psychotic disorders | 0.010 | 0.162 | 0.952 | 0.148 | 0.118 | 0.211 | 0.156 | 0.150 | 0.302 |
| | Mood disorders | -0.018 | 0.049 | 0.708 | 0.008 | 0.036 | 0.818 | 0.014 | 0.044 | 0.758 |
| | Anxiety- and stress-related disorders | -0.004 | 0.002 | 0.031 | -0.003 | 0.001 | 0.032 | -0.002 | 0.002 | 0.109 |
| | Behavioral syndromes with physical components | 0.043 | 0.074 | 0.565 | 0.009 | 0.055 | 0.868 | 0.061 | 0.076 | 0.421 |

B is the Mendelian randomization effect estimate from the corresponding method (MR-Egger, weighted median, or weighted mode).

SE = standard error. All analyses are two-sample MR using summary-level GWAS data for gynecologic disorders and psychiatric outcomes.

**Supplemental Table S12: Heterogeneity statistics (Cochran's Q) for two-sample Mendelian randomization analyses (IVW and MR-Egger) estimating the effect of genetic liability to gynecologic disorders on psychiatric outcomes.**

| Gynecologic disorder | Psychiatric Outcome | MR-Egger | | | Inverse variance weighted | | |
|---|---|---|---|---|---|---|---|
| | | Q | df | p-value | Q | df | p-value |
| Endometriosis | **Schizophrenia & other psychotic disorders** | 26.058 | 28.000 | 0.570 | 27.085 | 29.000 | 0.567 |
| | **Mood disorders** | 34.556 | 25.000 | 0.097 | 35.782 | 26.000 | 0.096 |
| | **Anxiety- and stress-related disorders** | 51.036 | 27.000 | 0.003 | 51.372 | 28.000 | 0.005 |
| | **Behavioral syndromes with physical components** | 27.667 | 28.000 | 0.482 | 28.086 | 29.000 | 0.513 |
| PCOS | **Schizophrenia & other psychotic disorders** | 9.644 | 6.000 | 0.140 | 9.833 | 7.000 | 0.198 |
| | **Mood disorders** | 2.759 | 7.000 | 0.906 | 2.814 | 8.000 | 0.945 |
| | **Anxiety- and stress-related disorders** | 7.150 | 6.000 | 0.307 | 7.218 | 7.000 | 0.407 |
| | **Behavioral syndromes with physical components** | 6.402 | 6.000 | 0.380 | 7.870 | 7.000 | 0.344 |
| Fibroids | **Schizophrenia & other psychotic disorders** | 75.813 | 66.000 | 0.191 | 75.920 | 67.000 | 0.213 |
| | **Mood disorders** | 56.523 | 65.000 | 0.764 | 56.920 | 66.000 | 0.780 |
| | **Anxiety- and stress-related disorders** | 56.261 | 64.000 | 0.743 | 57.659 | 65.000 | 0.729 |
| | **Behavioral syndromes with physical components** | 51.877 | 66.000 | 0.898 | 52.865 | 67.000 | 0.896 |

Q = Cochran's Q statistic for heterogeneity across SNP-specific causal estimates.

df = degrees of freedom for Q; p-value = p-value for the heterogeneity test.

MR-Egger and inverse variance weighted (IVW) models were implemented in a two-sample Mendelian randomization framework using summary-level GWAS data.

**Supplemental Table S13: MR-Egger intercept tests for horizontal pleiotropy in two-sample Mendelian randomization analyses estimating the effect of genetic liability to gynecologic disorders on psychiatric outcomes.**

| Gynecologic disorder | Psychiatric Outcome | MR-Egger Intercept | SE | p-value |
|---|---|---|---|---|
| Endometriosis | Schizophrenia & other psychotic disorders | 0.025 | 0.024 | 0.319 |
| | Mood disorders | -0.009 | 0.009 | 0.355 |
| | Anxiety- and stress-related disorders | 0.000 | 0.000 | 0.677 |
| | Behavioral syndromes with physical components | -0.008 | 0.012 | 0.523 |
| PCOS | Schizophrenia & other psychotic disorders | 0.024 | 0.070 | 0.743 |
| | Mood disorders | 0.004 | 0.017 | 0.821 |
| | Anxiety- and stress-related disorders | 0.000 | 0.001 | 0.820 |
| | Behavioral syndromes with physical components | 0.033 | 0.028 | 0.285 |
| Fibroids | Schizophrenia & other psychotic disorders | 0.004 | 0.014 | 0.761 |
| | Mood disorders | 0.003 | 0.004 | 0.531 |
| | Anxiety- and stress-related disorders | 0.000 | 0.000 | 0.241 |
| | Behavioral syndromes with physical components | -0.006 | 0.006 | 0.324 |

The MR-Egger intercept represents the average directional pleiotropic effect across genetic instruments.
A statistically significant intercept ($p < 0.05$) indicates evidence of directional horizontal pleiotropy.
SE = standard error.

**Supplemental Figure S3: Scatter plots of SNP effects on Endometriosis and psychiatric outcomes inverse-variance weighted, MR Egger, weighted median, and weighted mode estimates**

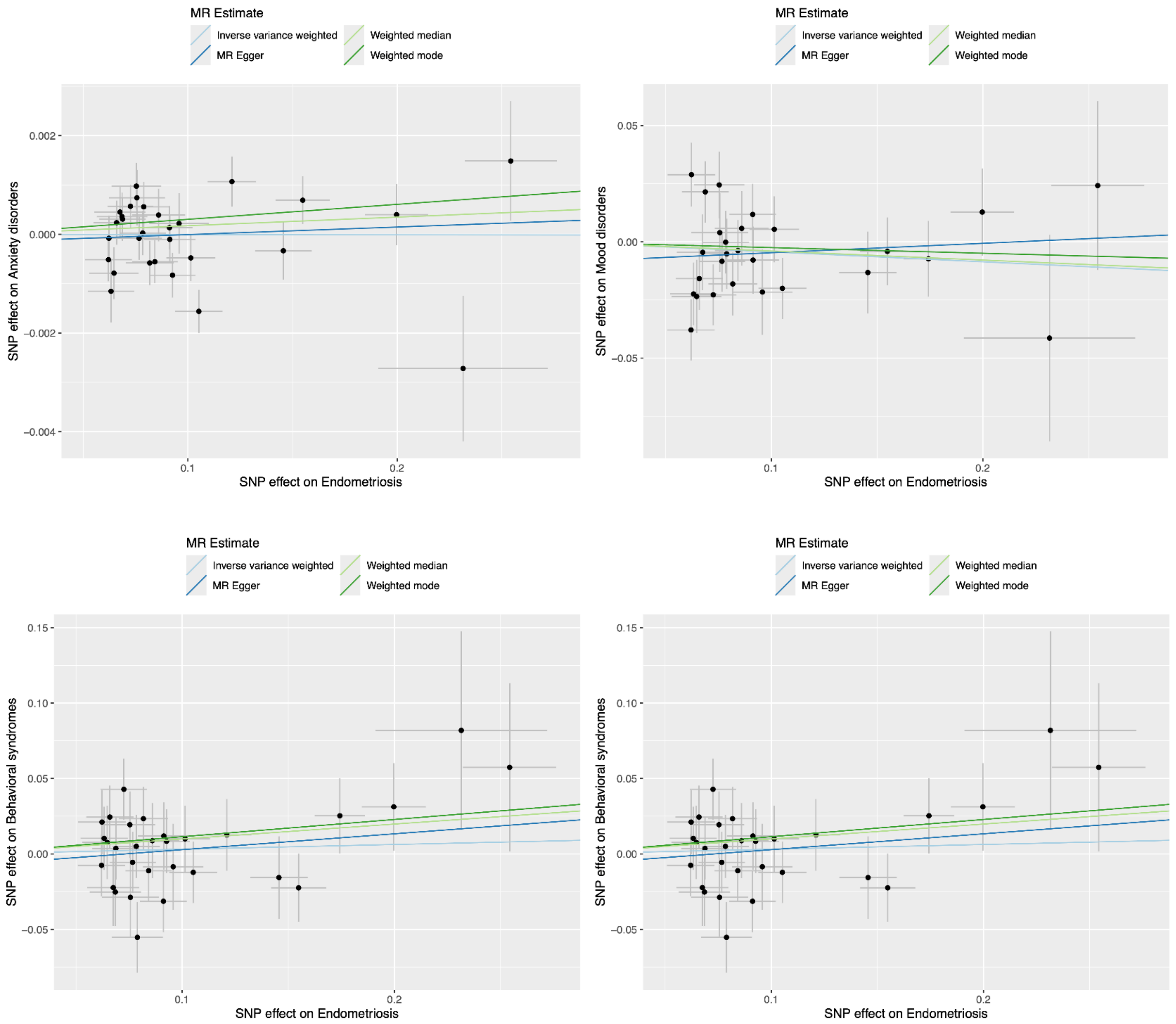

**Supplemental Figure S4: Scatter plots of SNP effects on PCOS and psychiatric outcomes I inverse-variance weighted, MR Egger, weighted median, and weighted mode estimates**

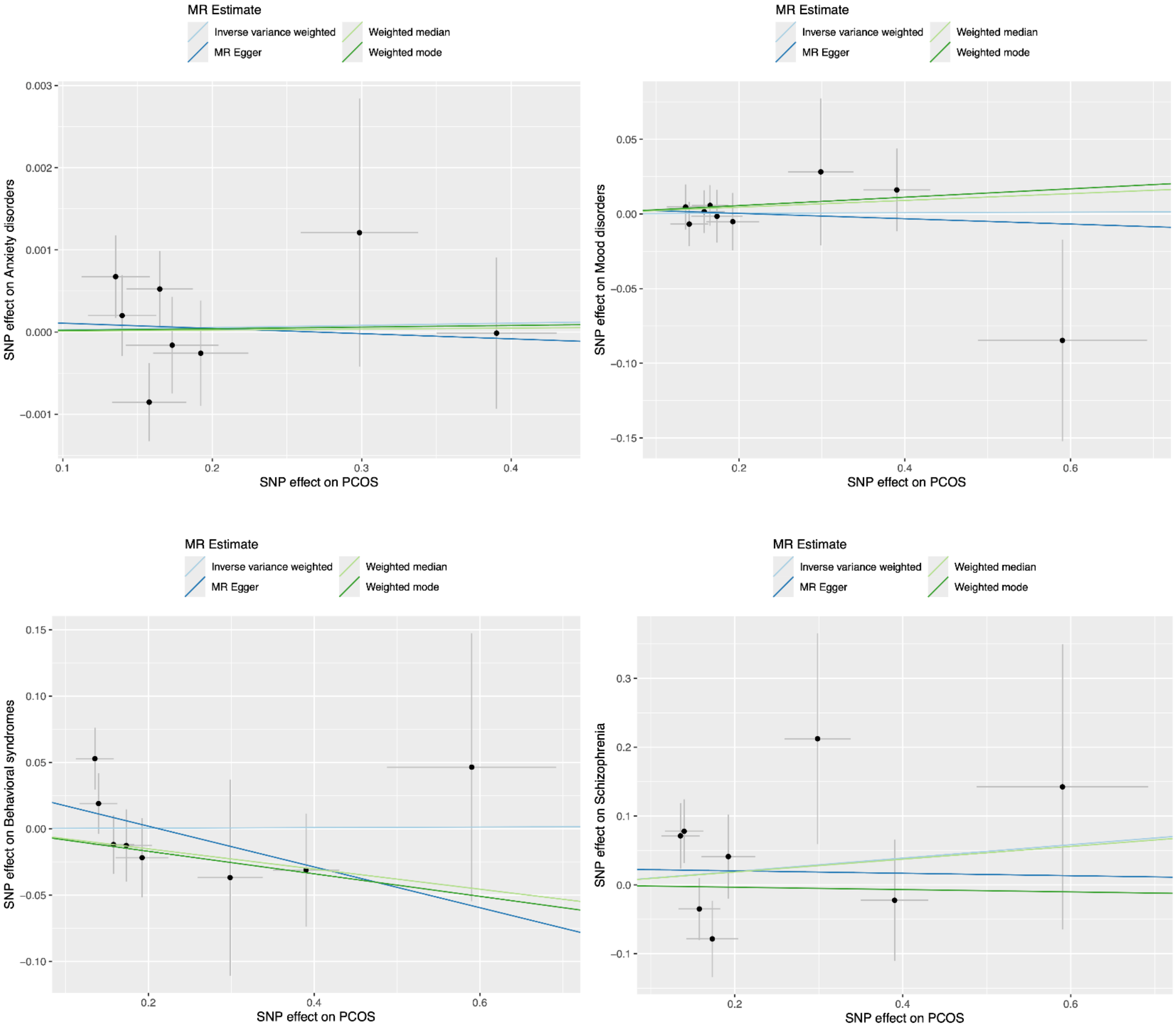

**Supplemental Figure S5: Scatter plots of SNP effects on Uterine Fibroids and psychiatric outcomes inverse-variance weighted, MR Egger, weighted median, and weighted mode estimates**

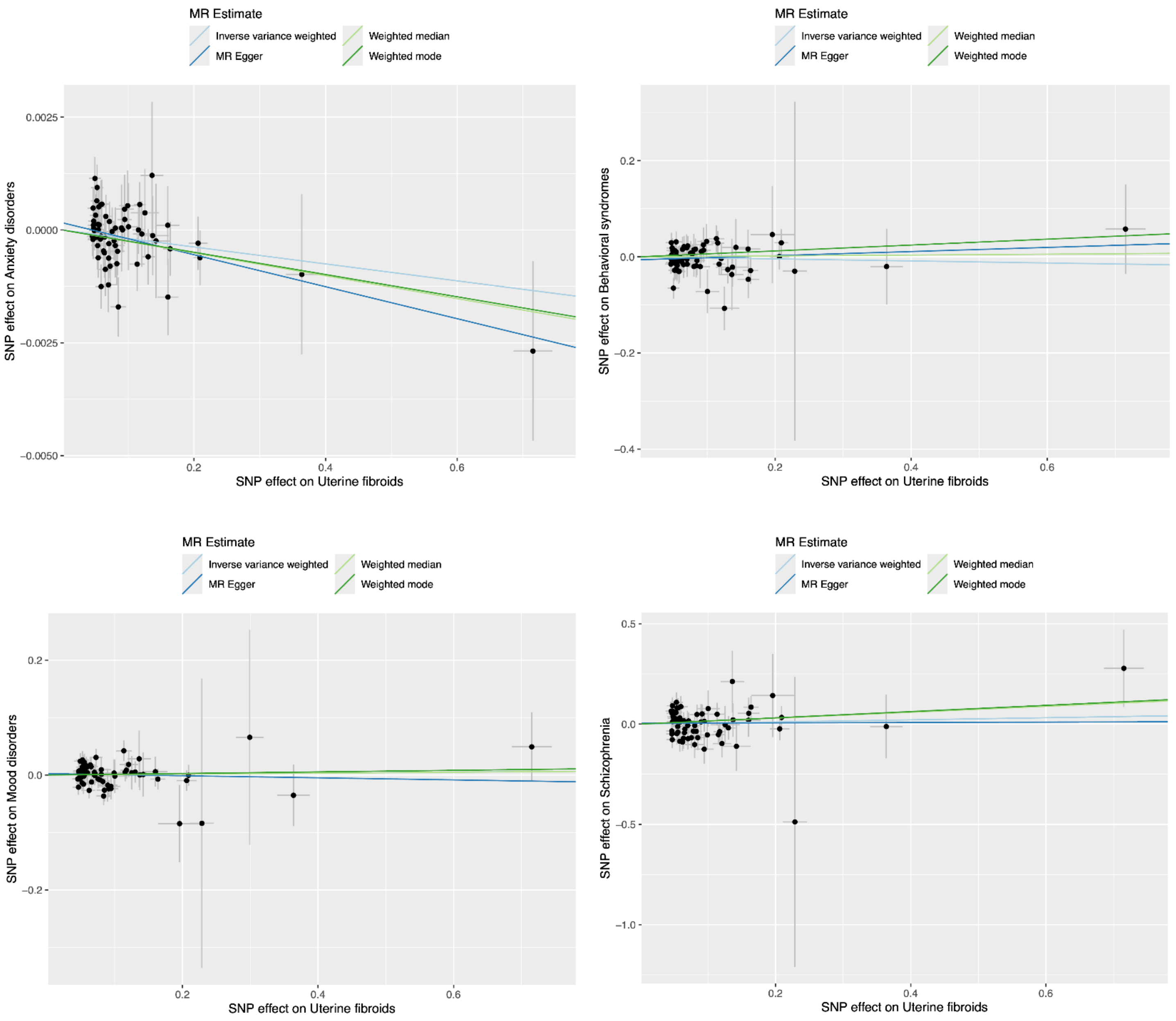

**Supplemental Table S14: Two-sample Mendelian Randomization analysis using inverse variance weighting testing psychiatric disorders as exposures and gynecologic disorders as outcomes.**

| Psychiatric Outcome | Gynecologic disorder | B | SE | p-value |
|---|---|---|---|---|
| **Schizophrenia & other psychotic disorders** | **Endometriosis** | 0.021 | 0.021 | 0.322 |
| | **PCOS** | 0.036 | 0.033 | 0.274 |
| | **Fibroids** | 0.027 | 0.013 | 0.047 |
| **Mood disorders** | **Endometriosis** | 0.021 | 0.041 | 0.612 |
| | **PCOS** | 0.028 | 0.081 | 0.727 |
| | **Fibroids** | 0.007 | 0.032 | 0.835 |
| **Anxiety- and stress-related disorders** | **Endometriosis** | 0.894 | 1.564 | 0.568 |
| | **PCOS** | -1.262 | 2.363 | 0.593 |
| | **Fibroids** | -0.671 | 1.153 | 0.560 |
| **Behavioral syndromes with physical components** | **Endometriosis** | 0.000 | 0.008 | 0.969 |
| | **PCOS** | -0.008 | 0.025 | 0.744 |
| | **Fibroids** | -0.002 | 0.007 | 0.727 |

B is the inverse-variance weighted two-sample Mendelian randomization effect estimate for the association between genetically predicted gynecologic disorders and psychiatric outcomes, expressed on the original GWAS scales of the exposure and outcome.
SE = standard error.

**Supplemental Table S15: Two-sample Mendelian Randomization analyses (MR-Egger, weighted median, and weighted mode) estimating the effect of genetic liability to psychiatric outcomes on gynecologic disorders.**

| Psychiatric Outcome | Gynecologic disorder | MR-Egger | | | Weighted median | | | Weighted mode | | |
|---|---|---|---|---|---|---|---|---|---|---|
| | | B | SE | p-value | B | SE | p-value | B | SE | p-value |
| Schizophrenia & other psychotic disorders | Endometriosis | 0.019 | 0.069 | 0.786 | 0.025 | 0.023 | 0.278 | 0.024 | 0.042 | 0.578 |
| | PCOS | 0.102 | 0.110 | 0.376 | 0.050 | 0.045 | 0.267 | 0.084 | 0.080 | 0.314 |
| | Fibroids | 0.031 | 0.044 | 0.501 | 0.021 | 0.018 | 0.247 | 0.019 | 0.045 | 0.687 |
| Mood disorders | Endometriosis | -0.095 | 0.096 | 0.333 | 0.021 | 0.056 | 0.710 | 0.036 | 0.217 | 0.869 |
| | PCOS | 0.058 | 0.217 | 0.793 | 0.029 | 0.112 | 0.794 | 0.021 | 0.453 | 0.963 |
| | Fibroids | 0.023 | 0.083 | 0.781 | 0.015 | 0.045 | 0.736 | 0.051 | 0.109 | 0.646 |
| Anxiety- and stress-related disorders | Endometriosis | -1.506 | 3.843 | 0.700 | 1.003 | 1.913 | 0.600 | 1.416 | 4.291 | 0.745 |
| | PCOS | -0.989 | 5.733 | 0.865 | -0.199 | 3.266 | 0.952 | 1.503 | 10.697 | 0.890 |
| | Fibroids | -0.885 | 2.980 | 0.770 | 0.277 | 1.493 | 0.853 | 0.080 | 5.588 | 0.989 |
| Behavioral syndromes with physical components | Endometriosis | 0.000 | 0.009 | 0.983 | 0.000 | 0.009 | 0.970 | 0.000 | 0.137 | 0.999 |
| | PCOS | 0.007 | 0.028 | 0.796 | 0.000 | 0.029 | 0.993 | 0.003 | 0.244 | 0.990 |
| | Fibroids | -0.004 | 0.008 | 0.607 | -0.004 | 0.007 | 0.560 | -0.003 | 0.114 | 0.976 |

B is the Mendelian randomization effect estimate from the corresponding method (MR-Egger, weighted median, or weighted mode).
SE = standard error. All analyses are two-sample MR using summary-level GWAS data for gynecologic disorders and psychiatric outcomes.

**Supplemental Table S16: Heterogeneity statistics (Cochran's Q) for two-sample Mendelian randomization analyses (IVW and MR-Egger) estimating the effect of genetic liability to gynecologic disorders on psychiatric outcomes.**

| Psychiatric Outcome | Gynecologic disorder | MR-Egger | | | Inverse variance weighted | | |
|---|---|---|---|---|---|---|---|
| | | Q | df | p-value | Q | df | p-value |
| Schizophrenia & other psychotic disorders | Endometriosis | 18.140 | 11.000 | 0.078 | 18.142 | 12.000 | 0.111 |
| | PCOS | 6.367 | 12.000 | 0.896 | 6.752 | 13.000 | 0.914 |
| | Fibroids | 15.583 | 13.000 | 0.272 | 15.594 | 14.000 | 0.339 |
| Mood disorders | Endometriosis | 21.063 | 18.000 | 0.276 | 23.154 | 19.000 | 0.231 |
| | PCOS | 21.757 | 18.000 | 0.243 | 21.783 | 19.000 | 0.295 |
| | Fibroids | 21.615 | 17.000 | 0.200 | 21.676 | 18.000 | 0.247 |
| Anxiety- and stress-related disorders | Endometriosis | 28.427 | 17.000 | 0.040 | 29.213 | 18.000 | 0.046 |
| | PCOS | 13.690 | 18.000 | 0.749 | 13.693 | 19.000 | 0.801 |
| | Fibroids | 26.895 | 18.000 | 0.081 | 26.904 | 19.000 | 0.107 |
| Behavioral syndromes with physical components | Endometriosis | 6.405 | 7.000 | 0.493 | 6.406 | 8.000 | 0.602 |
| | PCOS | 3.982 | 7.000 | 0.782 | 5.698 | 8.000 | 0.681 |
| | Fibroids | 8.542 | 8.000 | 0.382 | 8.867 | 9.000 | 0.450 |

Q = Cochran's Q statistic for heterogeneity across SNP-specific causal estimates.
df = degrees of freedom for Q; p-value = p-value for the heterogeneity test.
MR-Egger and inverse variance weighted (IVW) models were implemented in a two-sample Mendelian randomization framework using summary-level GWAS data.

**Supplemental Table S17: MR-Egger intercept tests for horizontal pleiotropy in two-sample Mendelian randomization analyses estimating the effect of genetic liability to psychiatric outcomes on gynecologic disorders.**

| Psychiatric Outcome | Gynecologic disorder | MR-Egger Intercept | SE | p-value |
|---|---|---|---|---|
| **Schizophrenia & other psychotic disorders** | **Endometriosis** | 0.000 | 0.018 | 0.978 |
| | **PCOS** | -0.018 | 0.029 | 0.547 |
| | **Fibroids** | -0.001 | 0.012 | 0.926 |
| **Mood disorders** | **Endometriosis** | 0.011 | 0.008 | 0.198 |
| | **PCOS** | -0.003 | 0.018 | 0.885 |
| | **Fibroids** | -0.002 | 0.007 | 0.829 |
| **Anxiety- and stress-related disorders** | **Endometriosis** | 0.008 | 0.012 | 0.502 |
| | **PCOS** | -0.001 | 0.018 | 0.959 |
| | **Fibroids** | 0.001 | 0.009 | 0.939 |
| **Behavioral syndromes with physical components** | **Endometriosis** | 0.000 | 0.007 | 0.976 |
| | **PCOS** | -0.019 | 0.014 | 0.232 |
| | **Fibroids** | 0.003 | 0.005 | 0.596 |

The MR-Egger intercept represents the average directional pleiotropic effect across genetic instruments.
A statistically significant intercept ($p < 0.05$) indicates evidence of directional horizontal pleiotropy.
SE = standard error.

**Supplemental Figure S6. Scatter plots of SNP effects on Schizophrenia and gynecologic disorders with inverse-variance weighted, MR Egger, weighted median, and weighted mode estimates.**

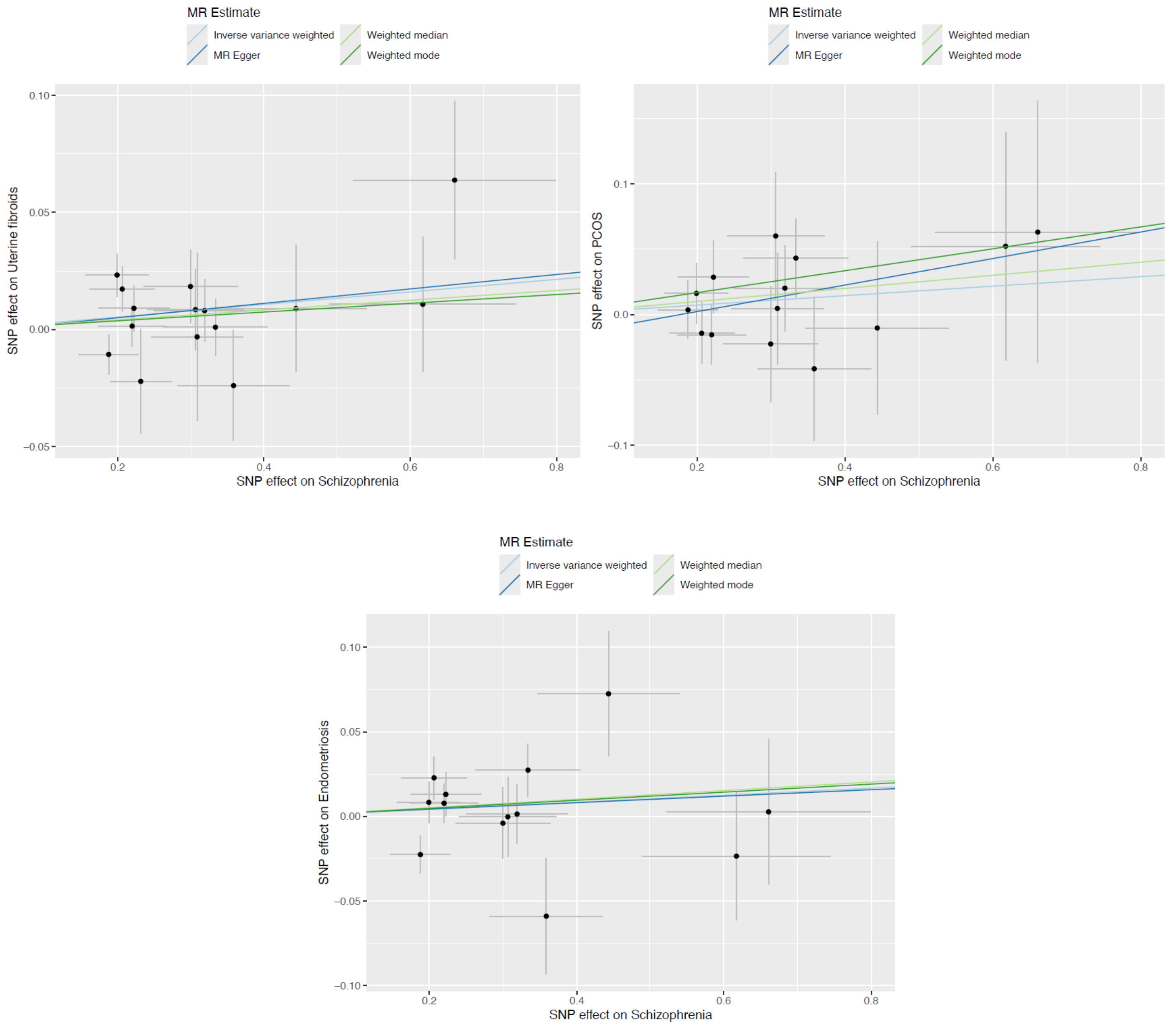

**Supplemental Figure S7. Scatter plots of SNP effects on Mood and gynecologic disorders with inverse-variance weighted, MR Egger, weighted median, and weighted mode estimates.**

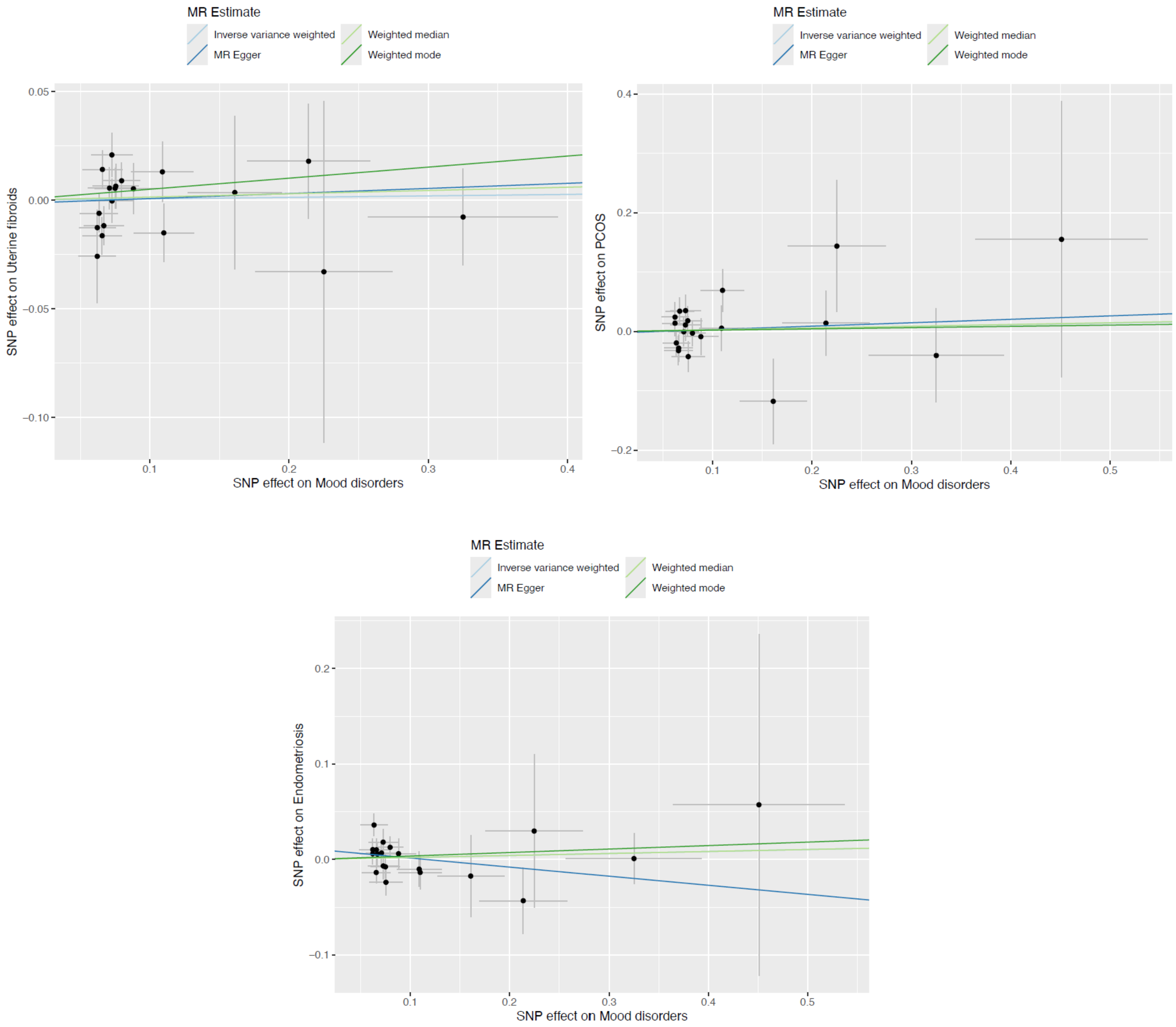

**Supplemental Figure S8. Scatter plots of SNP effects on Anxiety/ stress-related and gynecologic disorders with inverse-variance weighted, MR Egger, weighted median, and weighted mode estimates.**

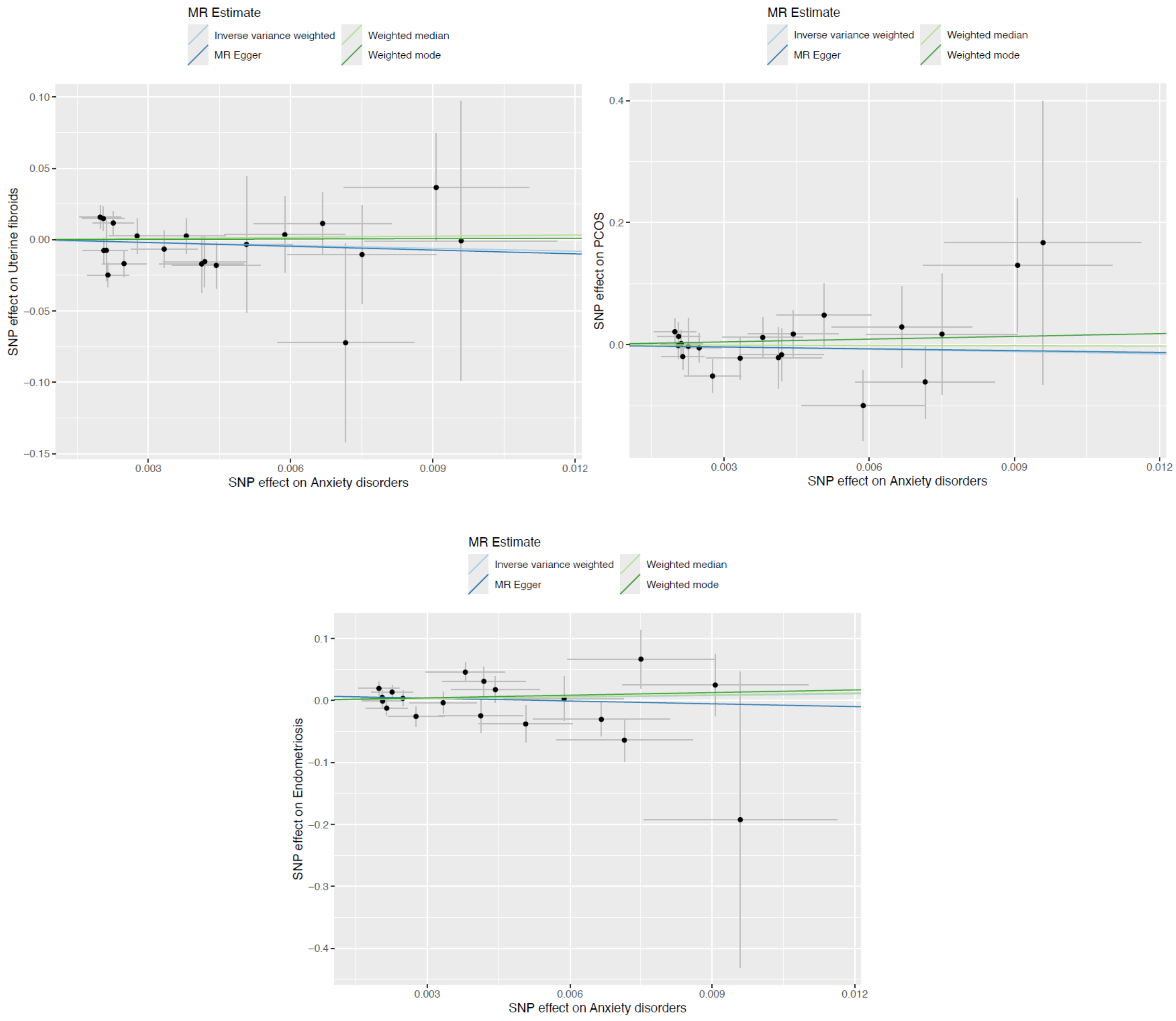

**Supplemental Figure S9. Scatter plots of SNP effects on Behavioral syndromes and gynecologic disorders with inverse-variance weighted, MR Egger, weighted median, and weighted mode estimates.**

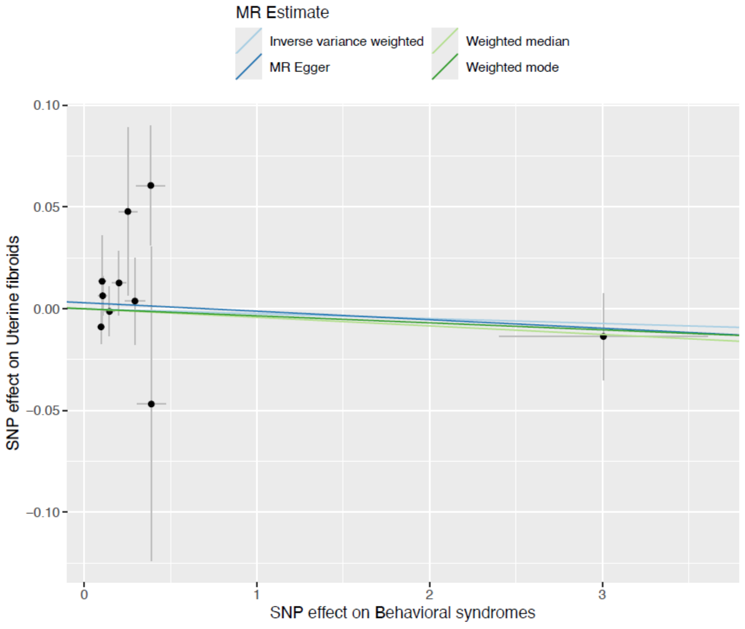


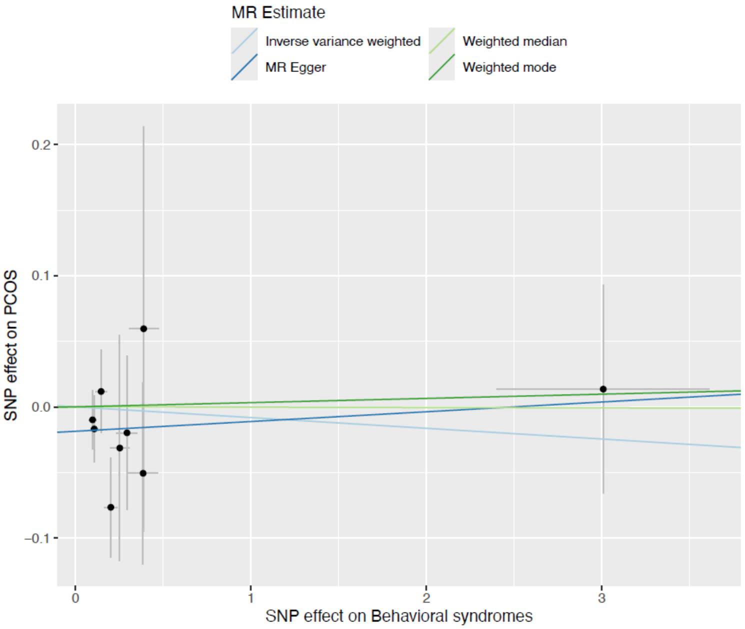


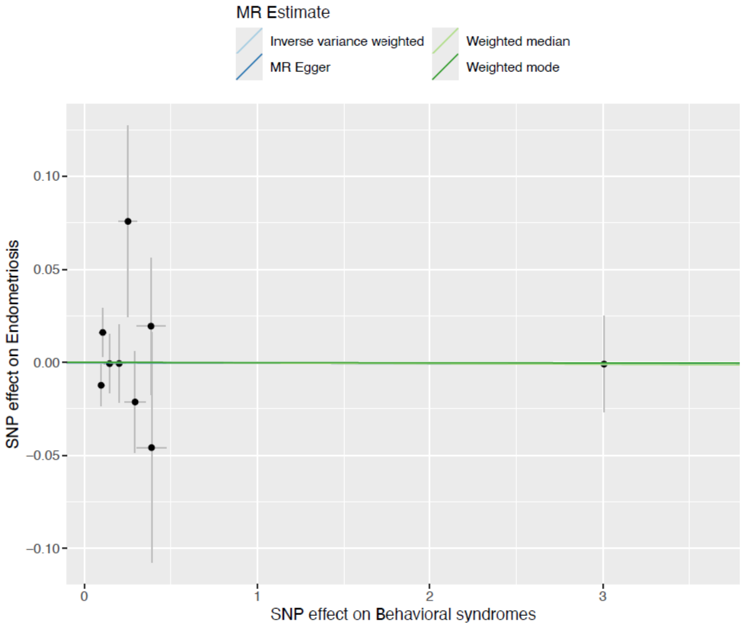
